# Multivariate Models using quantitative CT assessment for prediction of outcome after Subarachnoid Hemorrhage

**DOI:** 10.1101/2024.11.26.24317478

**Authors:** Marcin Balcerzyk, Juan José Egea-Guerrero, Gabriel Cepeda-Carrión, Ángel Parrado-Gallego, Zaida Ruiz de Azúa López, Ángel Vilches-Arenas, Javier Rodriguez-Santero, Enrique de Vega-Ríos, Manuel Quintana-Diaz, Soledad Perez-Sanchez, Julene Argaluza-Escudero, R. Loch Macdonald

## Abstract

**Objective:** To diagnose if a patient has subarachnoid hemorrhage (SAH) and to predict if he/she will develop angiographic vasospasm/delayed cerebral ischemia (DCI) or will die using only admission cranial computed tomography (CT) and World Federation of Neurological Surgeons (WFNS) or Hunt-Hess clinical grades.

**Methods:** We developed a semiautomatic method of quantification of SAH volume, surface, sphericity, fractal dimension, and other parameters by retrospectively analyzing admission CT scans of 127 SAH patients. A multivariable model to predict the development of vasospasm/DCI and death was fitted by combining CT and admission neurological grade with derived data. It was also developed and then validated with bootstrapping and with a separate group of patients.

**Results:** The diagnosis of SAH can be given by surface to volume ratio of the 60-80 HU volume of interest for blood AUC 0.98 (95% CI 0.95-1), hemorrhage volume 0.92 (95% CI 0.85-0.99), sphericity 0.87 (95% CI 0.73-1) and surface area 0.75 (95% CI 0.56-0.94). For vasospasm/DCI prediction AUC of biomarker panel was 0.731 (95%CI 0.626-0.836) which was significantly different from random (p<0.001) while Hunt-Hess gave 0.653 (95%CI 0.535-0.772). We established one individual image derived predictor of death by the surface to volume ratio at the level of AUC (Area under the curve) of 0.777 (95% CI 0.667 - 0.886), while a biomarker panel gave the value of AUC of 0.857 (95%CI 0.769-0.944). These two biomarkers allow the correct prediction of the death of nine of ten patients with 36% and 59% probability, respectively.

**Conclusions:** The model allows rapid, objective, and quantitative assessment of the risk of vasospasm/DCI and death in SAH patients. Both assessments, being fully automatic, can be easily introduced to CT scanner reconstruction software.

**Key message:**

- **What is already known on this topic** – Subarachnoid hemorrhage (SAH) has an incidence of 7.9 per 100 000 inhabitants per year and morbidity of almost 90%. It is typically diagnosed with a cranial CT-scan by a radiologist. The amount of blood on the CT scan is assessed qualitatively using the Fisher score and clinical evaluation through the WFNS and Hunt-Hess scales. The objective was to establish some unitary predictor for diagnosis and biomarker model predictors of death and vasospasm after distinguishing from control cases.
- **What this study adds** – Contrary to common sense knowledge the most significant factor in increasing of intra-hospital death probability is not the hemorrhage volume, but its surface-to-volume ratio derived from the image volume-of-interest delineated at 60-80 HU.
- **How this study might affect research, practice or policy** - This model could help physicians to take decisions on redirecting the patient to more monitored hospital areas, anticipate possibilities of complications and apply endovascular treatment with calcium antagonists or balloon angioplasty.

## 1 Introduction

Subarachnoid hemorrhage (SAH) affects about 7.9 (95% CI, 6.9-9.0) per 100,000 people per year worldwide and is associated with high morbidity and mortality^2^. It is typically diagnosed with computed tomography (CT) done in the emergency department. The severity of the SAH is assessed through clinical evaluation (e.g. Hunt-Hess scale or World Federation Neurological Surgeons (WFNS) score) and CT Scan information, such as the Fisher^3^, modified Fisher^4^ or Hijdra^5^ scales. For patients who survive the initial SAH, angiographic vasospasm/DCI^6^ is a frequent complication which can cause additional disability and even death usually between 4 and 14 days after the SAH.

If the CT-scan is not done within 6 hours or diagnosis is missed by doctors the outcome is poorer. On the other hand, if a diagnosis is more accurate one can avoid a lumbar puncture^7^. Whatever the timing and quality of the diagnosis is, the prognostic of the outcome is not clear. In recent years several single biomarkers have been proposed for the outcome in SAH ^8–17^.

In addition to its role in diagnosis of SAH, there is a need for more reliable or accurate CT scan interpretation since it is a key predictor of vasospasm/DCI and a prognostic factor for outcome after SAH. Studies have reported quantitative measures of SAH volumes, but the methods used have been time-consuming, and not automated. The prognostics are done usually for poor outcomes or the occurrence of vasospasm/DCI. Poor outcome is based on emergency head CT scan and/or some clinical data ^18–20^, while the most successful vasospasm/DCI prediction was done on perfusion CT scans acquired outside of the emergency department and several days after admission^21–23^. In general, the volumetric evaluation of the bleeding is manual and slow, taking at best 20 min^24^ or offers no other information than hemorrhage volume and distribution^25^ ^26^. The volume of SAH (usually in 60-80 HU range, but see^26^) is widely considered as the best indicator of poor outcome (for SAH see Ref.^19^ ^24^ ^27^ and for intracerebral hemorrhage see Ref. ^28^ ^29^).

Herein we present a semiautomatic method of differentiating between normal CT and CT of patients presenting with SAH as well as a semiautomatic prognostic method of forecasting of the occurrence of vasospasm/DCI and intra-hospital death based on emergency department CT and routinely and immediately available clinical data. The study is a development project with both bootstrap, hold-out sampling, and internal validation. The method is based on logistic regression. The semiautomatic evaluation takes about 1-3 minutes on a fast computer. The report was prepared using STROBE^30^ and TRIPOD^31^ guidelines.

## 2 Methods

### 2.1 Study design and participants

We performed a retrospective observational cohort study of patients who were admitted to ICU after Severe SAH. The study was performed between 2012 and 2014 in Hospital Universitario Virgen del Rocio (HUVR, Sevilla, Spain) and in Hospital Universitario La Paz, Madrid, Spain (HULP). The study flow chart is shown in Suppl. Figure 1. The dataset contained 136 subjects: 127 SAH confirmed patients (96 from HUVR and 31 from HULP) and 9 controls from HUVR. Controls were patients with suspected cranial trauma who were later not diagnosed with any abnormalities.

There were another 13 control patients in HUVR whose data was used for elaboration of the model, but not in the final development. 34 of 127 patients suffered intra-hospital-death and 76 a vasospasm. 81 patients were women and 46 men. Clinical data included age, sex, admission neurological grade (Hunt-Hess and, WFNS grades), and modified Fisher scale data as determined in Supplementary Section 8.4. The dependent data were development of vasospasm/DCI, and an outcome at the end of hospitalization (death/alive). The inclusion criteria were subarachnoid hemorrhage diagnosis of spontaneous SAH and availability of CT scans from the emergency department up to two hours from arrival to the hospital. The exclusion criteria were incomplete head CT scan, movement during the CT scan, the presence of metallic artifacts in the image (e.g. external ventricular drainage or endovascular coils). If there was no information about vasospasm/DCI or death, the data were treated as missing (n=3). All patients had data record of occurrence or not of intra-hospital death.

The patient’s level of consciousness was assessed using the Glasgow coma scale (GCS). It was determined on admission after hemodynamic and metabolic resuscitation, ventricular drainage, if necessary, exclusion of effects of sedatives and drugs, and then every 24 hours.

The clinical grade of the patient was determined based on the Hunt and Hess (HH)^32^ without adjustment for severe illness and the World Federation of Neurological Surgeons (WFNS) scales^33^. The patient details, treatment and study flow chart are described in Supplementary Section 8.1 and approval in Supplementary Section 8.2.

### 2.2 Head CT scan

CT images were acquired with various protocols purposely to test the efficiency of the procedures. The CT systems used were Toshiba Aquilon, GE LightSpeed 16, and Siemens Sensation 16. The image resolution was from 0.3 mm to 5 mm in the head-feet axis of the patient and from 0.4 mm to 0.5 mm in sagittal and coronal axes.

### 2.3 Image processing

Initially, the CT images acquired at admission were examined for errors including patient movement, incomplete skull scan, and metal artifacts, and these images were excluded. Only patients with known clinical parameters were examined (except for three patients with unknown data on vasospasm).

Briefly summarizing the procedure, the images were loaded to PMOD 4.3^34^ and examined for appropriate quality, and saved in the PMOD database. After examining several sample patients and some controls, the workflow was set in Batch processing of PMOD. The details are described in Supplementary Section 8.3

### 2.4 Statistical analysis

The clinical independent data included age, sex, Hunt-Hess, WFNS, and modified Fisher scale data. The dependent data were subsequent development of vasospasm/DCI or not, and an outcome at the end of hospitalization (death/alive).

The primary outcome was the diagnosis of vasospasm/DCI during the first admission. The secondary outcome measure was death during admission. No actions to the blind assessment of the outcome to be predicted were taken.

All statistical analysis was done in Excel 2019, SPSS version 23 to 25 (IBM, New York), STATA (Stata corp., College Station) and R version 3.6.1 and 4.4.1 with pROC and Rcmdr packages. The significance level was set at 0.05. The Area under the curve/Receiver operating characteristics (AUC/ROC) were used to examine if individual image derived or clinical variables explained in-hospital death or vasospasm/DCI. The optimum cutoff value was calculated in pROC with Youden statistics. Another characteristic is partial Area Under the Curve (pAUC) which indicates the quality of a biomarker at high specificity^35^. We selected the pAUC for the region of 100-90% specificity (i.e. initial 0.1 of the 1-*specificity* curve). Note that for a perfect case of AUC=100%, the described pAUC would be only 10%. The selection of 90% specificity was driven by the necessity to correctly predict the outcome for nine out of ten cases. The details of statistical analysis are described in Supplementary Section 8.4.

## 3 Results

The workflow in our study is summarized in Figure 1. Patient characteristics are presented in Table 1. The typical patient images, data, and evaluation are shown in Supplementary Section 8.5. There was no significant difference between groups from both hospitals except for age. The values of the variables and the descriptive statistics are shown in Suppl. Table 1.

**Figure 1.**
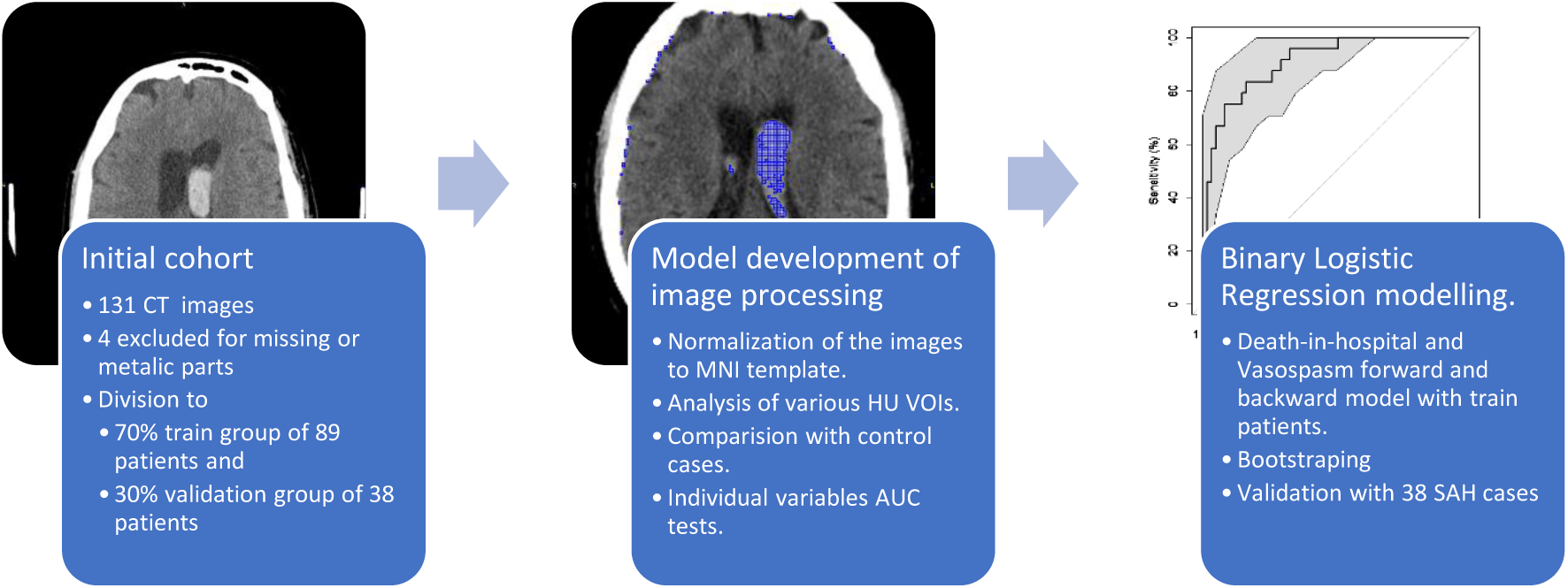
Workflow of the study

**Table 1.** Patientś characteristics. . The patients whose images were used in the development of the model.

| Characteristics | All patients | Death in the hospital = TRUE | Vasospasm/DCI = TRUE* |
| --- | --- | --- | --- |
| Age, yrs. | 55.0 | 60.1 | 52.3 |
| Men, yrs. (n=33) | 54.2 | 58.8 | 53.7 |
| Women, yrs., (n=52) | 55.4 | 61.1 | 51.6 |
|  | *Count* | *Count* | *Count* |
| n | 127 | 34 | 48 |
| Hunt-Hess value |  |  |  |
| 1 | 15 | 2 | 2 |
| 2 | 27 | 2 | 8 |
| 3 | 19 | 5 | 9 |
| 4 | 32 | 10 | 16 |
| 5 | 34 | 15 | 13 |
| WFNS values | *Count* | *Count* | *Count* |
| 1 | 19 | 1 | 4 |
| 2 | 27 | 4 | 10 |
| 3 | 11 | 2 | 4 |
| 4 | 39 | 13 | 18 |
| 5 | 31 | 14 | 12 |
| Modified Fisher scale value | *Count* | *Count* | *Count* |
| 1 | 2 | 0 | 0 |
| 2 | 9 | 1 | 1 |
| 3 | 23 | 5 | 14 |
| 4 | 93 | 28 | 33 |

\* Data of three patients on vasospasm/DCI were missing and were treated as such.

### 3.1 Comparison between SAH patients and controls and diagnostic capabilities

Table 2 shows that the automated protocol for the estimation of intracranial bleeding makes it possible to differentiate between patients without intracranial hemorrhage (controls) and patients with SAH. Namely, the volume of the hemorrhage, the surface area of the hemorrhage, and surface to volume ratio are all significantly different between SAH patients and controls. As shown in Figure 2, surface to volume ratio has reached AUC of 0.98 (95% CI 0.95-1.00) followed by volume at AUC of 0.92 (0.85-0.99) and surface with AUC of 0.75 (0.56-094). Note that the effect size is about 0.9 and for the surface to volume ratio is more than 2.

**Figure 2.**
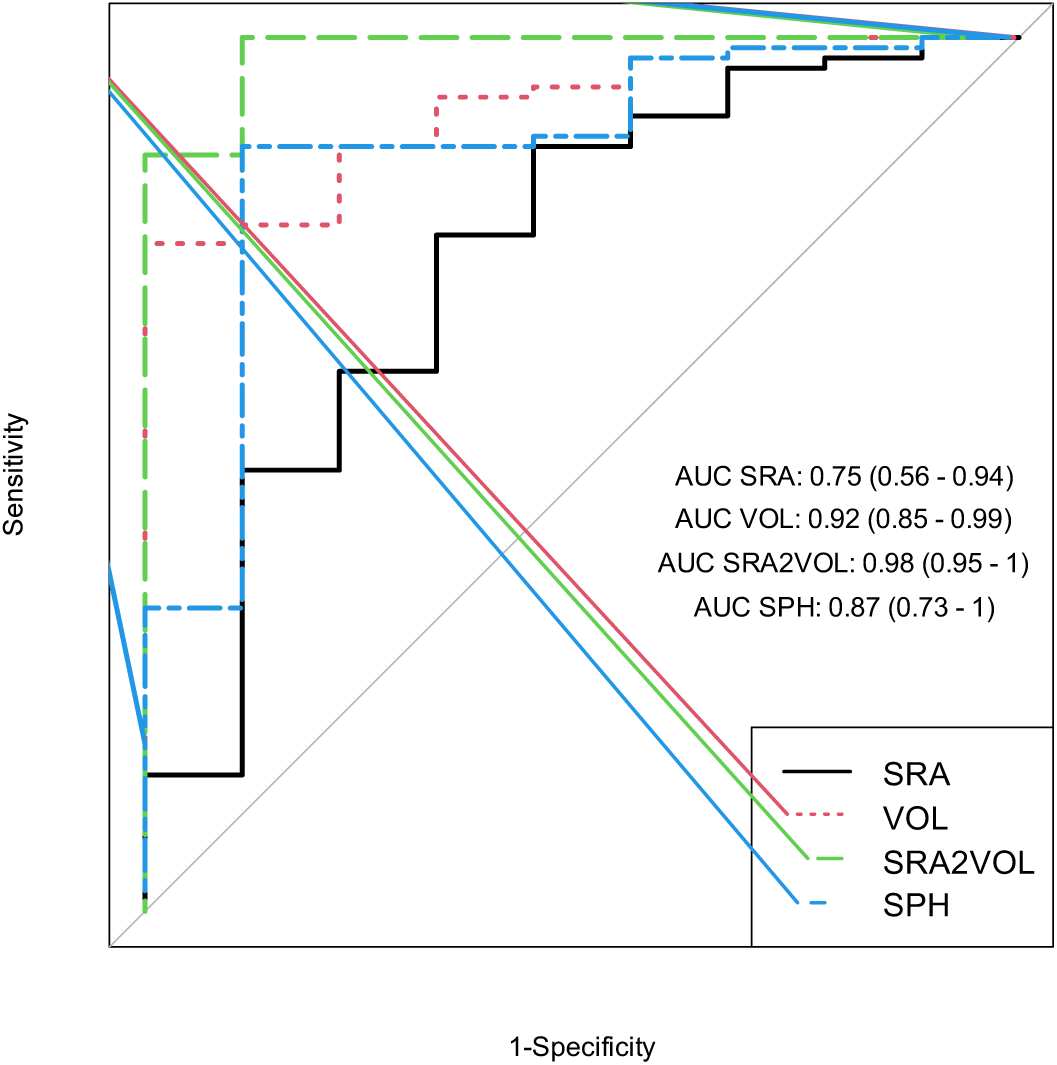
AUC/ROC curves for diagnostics of SAH with three image derived biomarkers. SRA signifies surface area of VOI at 60-80 HU in cm^2^, VOL volume in cm^3^ (mL), SPH is dimensionless, SRA2VOL is the ratio of SRA to VOL expressed in cm^-1^.

**Table 2.** Difference between healthy subjects (controls) and SAH patients. . Ranges in parentheses are 95% confidence intervals, otherwise they are standard deviations. One value in parentheses corresponds to standard deviation. P was calculated with Wilcoxon test, as for controls the distribution was not normal. d is the effect size for which we used standard deviations calculated with Hedges correction. There were 9 controls and 89 SAH patients. There were 38 validation SAH patients. There was no difference between train and validation groups. The significance of Ctrl-Train and Ctrl-Valid comparison was the same. More detailed characteristics are provided in Suppl. Table 7 in Section 8.6

| Patients |  |  |  |  |  |  |  |  |  |
| --- | --- | --- | --- | --- | --- | --- | --- | --- | --- |
| CONTROL | SAH | p | d | AUC | Threshold | Sensitivity, % | Specificity, % | PPV, % | NPV, % |
| **The volume of the hemorrhage, mL** |  |  |  |  |  |  |  |  |  |
| 6.32<br>(3.35) | 22.46<br>(17.66) | 0.021 | 1.26 | 0.92<br>(0.85-0.99) | 11.4 | 100 | 88.9 | 70 | 0 |
| **The surface area of the hemorrhage, cm<sup>2</sup>** |  |  |  |  |  |  |  |  |  |
| 461 (215) | 679<br>(238) | 0.022 | 0.952 | 0.75<br>(0.56-0.94) | 532 | 77.5 | 66.7 | 76.9 | 4.2 |
| **The sphericity of the hemorrhage, dimensionless.** |  |  |  |  |  |  |  |  |  |
| 0.037<br>(0.007) | 0.054<br>(0.021) | <0.001 | 1.12 | 0.87<br>(0.73-1) | 0.0379 | 87.6 | 88.9 | 57.9 | 1.3 |
| **Surface area to volume ratio of the hemorrhage, cm<sup>-1</sup>** |  |  |  |  |  |  |  |  |  |
| 75.7 (8.5) | 39.6<br>(15) | <0.001 | 2.92 | 0.98<br>(0.95-1) | 69.7 | 100 | 88.95 | 0 | 1.1 |

### 3.2 Vasospasm/DCI characteristics

We examined single variable AUC/ROC (volume, surface area, sphericity, the diameter of the hemorrhage, fractal dimension, surface to volume ration, age, sex, HH, WFNS, and modified Fisher Scale, see Figure 3 and Suppl. Table 1) and a panel model to develop a predictor of vasospasm/DCI episode. No single parameter was a significant predictor in forward selection. There were n=3 patients for whom there were vasospasm data missing. Figure 3, Suppl. Table 1, Suppl. Table 2

**Figure 3.**
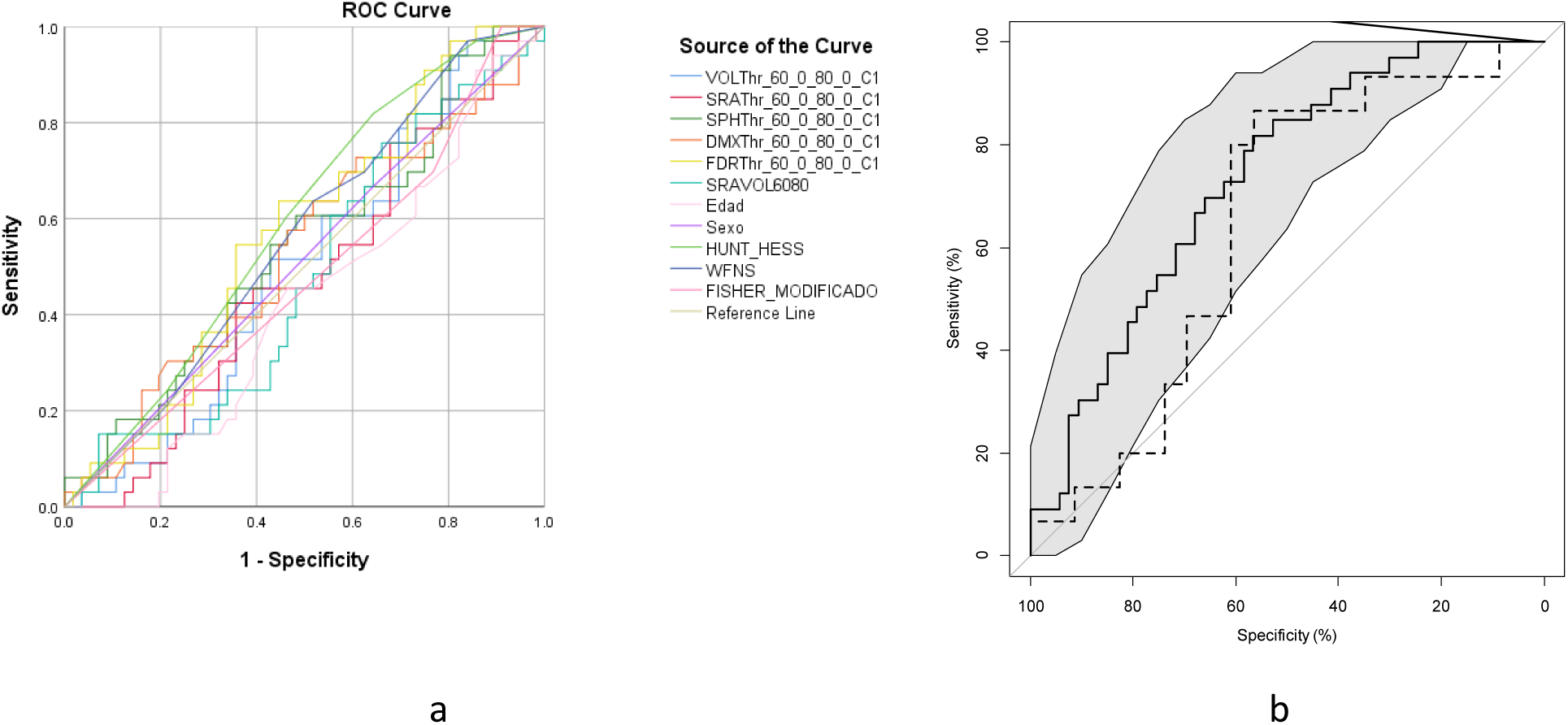
AUC/ROC curves for vasospasm/DCI. (a) AUC/ROC curves for vasospasm/DCI for individual variables. None is significant individually. (b) AUC/ROC 0.731 (CI 0.626 - 0.836) curve for Vasospasm/DCI model. The grey area corresponds to a 95% confidence interval. Note better model performance in upper (100-80%) specificity range, than individual predictors. The solid line is for training data, the dashed line is for validation data.

The Vasospasm/DCI backward model parameters are summarized in Suppl. Table 2 of which we mention the correct prediction (accuracy) of 67.1%. The details of the parameters used below are described in Supplementary Section 8.3 Image processing details.

In backward regression using all Volumes of Interest (VOI) and parameters mentioned in sections 2.2 and 8.3, gave the most significant results and highest AUC. The variables that entered the final model are fractal dimension (FDR, for 60-80 HU), volume (VOL, for 60-80 HU), maximum diameter (DMX, for 55-60 HU), and sphericity (SPH, for 60-80 HU), (although the last one not significant at p=0.101). The outcome of 81.1% (CI 72.6%-88.7%) of patients without vasospasm/DCI was predicted correctly (specificity) and only 45.5% (CI 31.7%-57.6%) with vasospasm/DCI was predicted correctly (sensitivity). The positive predictive value was 60.0%, the negative predictive value was 70.5% and accurately 67.4% (CI 56.9%-76.8%). The Nagelkerke R^2^ coefficient for this model is 0.23, which is a parameter that measures the fraction of variance described by the model.

AUC/ROC curve for Vasospasm/DCI model is shown with a central solid line and grayscale confidence interval area in Figure 3 (b) and the values are shown in Table 3. Note that AUC for the model is 0.731 (CI 0.626 - 0.836) and significant with pAUC of 1.4% out of 10%, way above the random level of 0.5%.

**Table 3.**
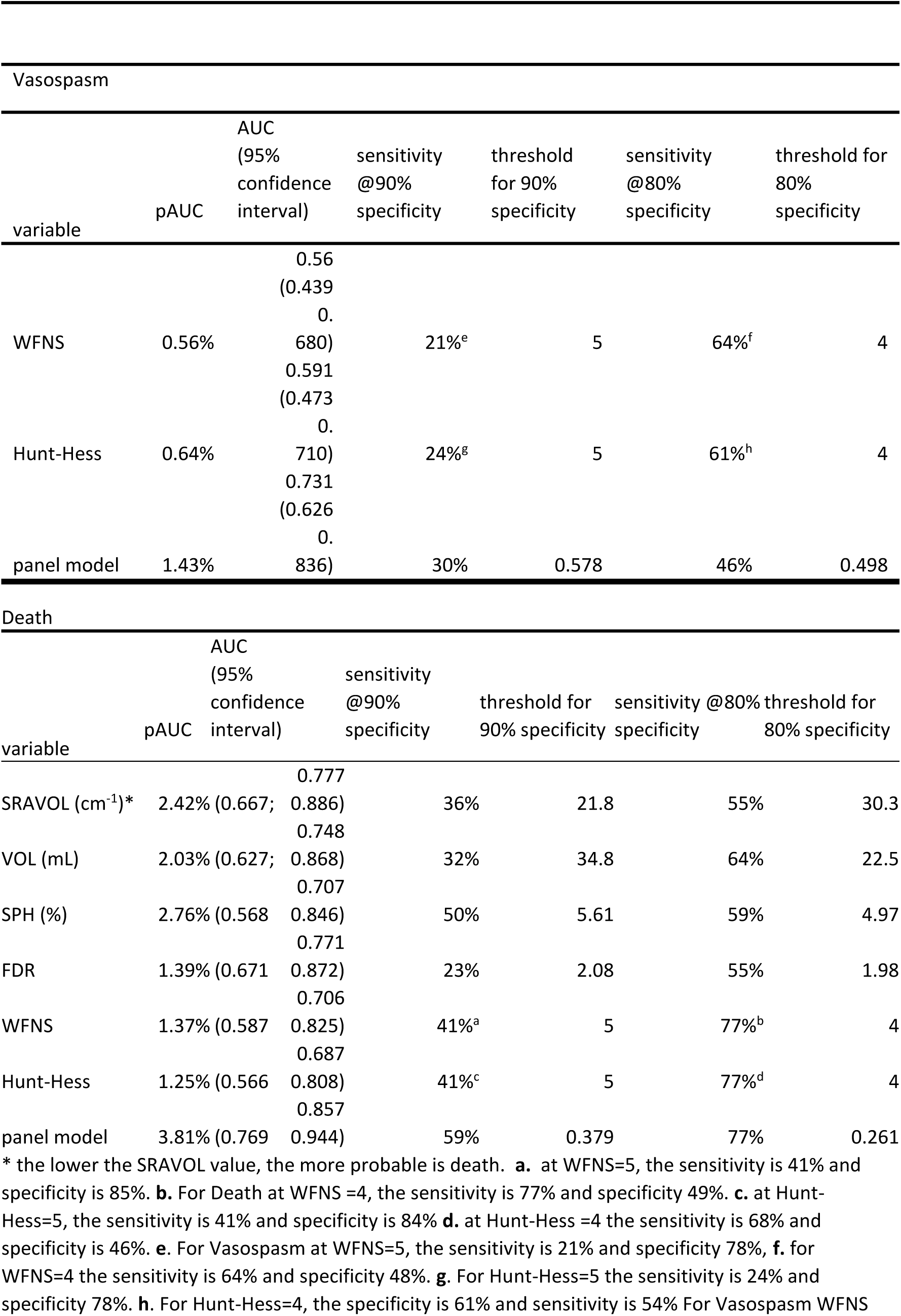
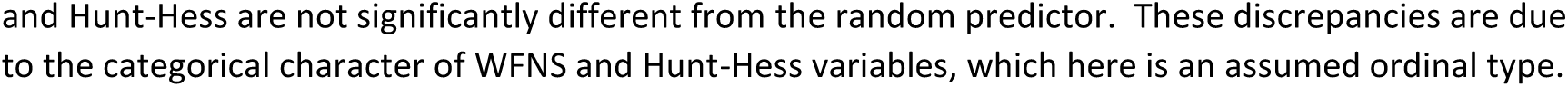
AUC/ROC with confidence intervals, pAUC sensitivities, and the threshold for individual parameters and panel model for intra-hospital death and vasospasm/DCI. Only statistically significant parameters are listed.

The step-change while removing parameters in a backward model was conducted and described in Suppl. Table 3 as the net reclassification index (NRI) to assess the decremental value of a variable on the prediction model. It was determined by the change in Youden’s J index, which reached 0.2847 in the final step.

### 3.3 In-hospital death characteristics

We examined single variables derived from an image or clinical ones as a predictor of in-hospital death. The results are shown in Figure 4 (a) and a Suppl. Table 1. All except one of the statistically significant image derived variables (surface to volume ratio, fractal dimension, volume, and sphericity) have higher AUC/ROC than statistically significant clinical variables of WFNS or Hunt-Hess scale.

**Figure 4.**
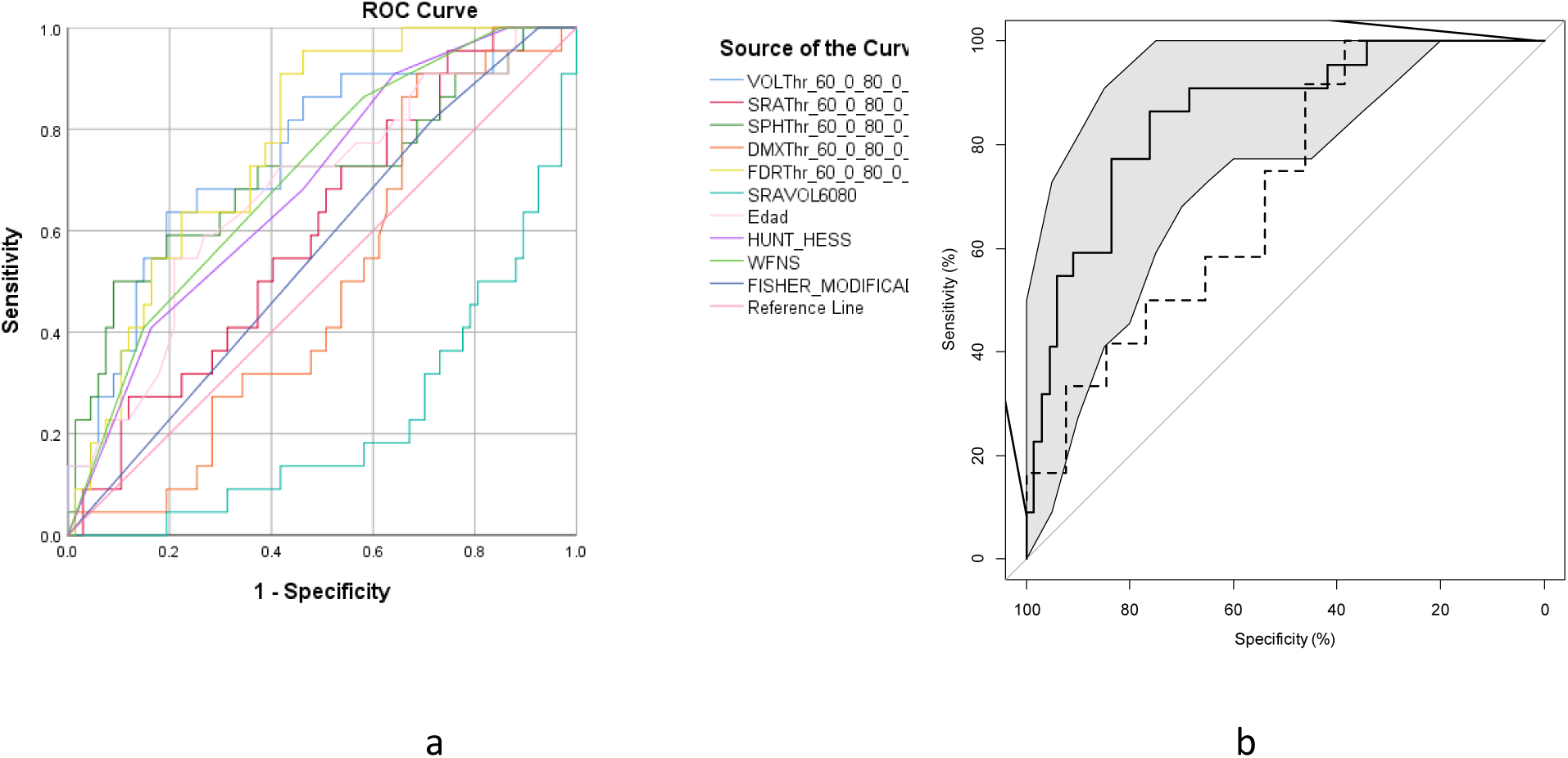
AUC/ROC curves for individual variables for Death (Exitus). (a) curves for individual variables for Death (Exitus) variable. Note that the SRAVOL variable has reverse direction than others, that means high-value prospects to survival. (b) AUC/ROC 0.857 (0.769 - 0.944) curve for the Death model. Grey range corresponds to a 95% confidence interval. The solid line is for training data, the dashed line is for validation data.

The data for AUC/ROC, and pAUC and threshold values for individual variables for 90% and 80% specificity are shown in Table 3. Surface to volume ratio of the VOI scored higher than volume in AUC, pAUC, and sensitivity at 90% specificity.

The regression iteration ended with the set of three variables: Surface-to-Volume, age, and sex with the statistical significance *p* increasing in the same order. Forward regression required four iterations; backward regression required eight iterations to complete. The Nagelkerke R^2^ coefficient for this model is 0.42.

The death model parameters and characteristics are summarized in Suppl. Table 4 Finally, the model resulted in accuracy of 83.1%, AUC of 0.857 (CI 0.769-0.944), pAUC of 3.81% out of maximum possible 10%. The model curve is shown in Figure 4 (b) with confidence intervals shown in the grey. The model correctly predicted 92.5% (CI 86.7%-96.8%) survival cases (specificity) and 54.5% (36.6%-67.5%) of death cases (sensitivity). The positive predictive value was 70.6%, the negative predictive value was 86.1% (CI 80.6%-90.1%), and accuracy 83.1% (CI 74.3%-89.6%). The Youden’s index reached 0.471 and the net reclassification index was expressed as change in Youden’s index. They are summarized in Suppl. Table 5.

### 3.4 Validation of the model

The model was validated on 38 patients selected randomly from the initial dataset, that were not used in training of the model. The validation and modeling groups did not differ in terms of age, sex, WFNS, HH, or hemorrhage volume determined from CT scan in 60-80 HU (data not shown). The accuracy of prediction is shown in Suppl. Table 6.

The ROC curves for validation data are shown as dashed lines in Figure 3 (b) and Figure 4 (b). The validation dashed curves are not significantly different from the solid model training curves as checked with a bootstrap test for two unpaired ROC curves.

The accuracy in the validation group was 57% for vasospasm/DCI and 73% for death. These values were close to the ones obtained in the modeling group, cf. Suppl. Table 2: 67% for vasospasm/DCI and Suppl. Table 4: 83% for death.

## 4 Discussion

### 4.1 Key novel findings

The novel findings of this study are that perhaps contrary to expectations, it was not the volume of the 60-80 HU VOI that was the best single predictor of Death, but Surface to volume ratio. The mechanistic interpretation of this finding may be that, for the values above the cut off value at 21.8 cm^-1^ for 90% specificity, patients with high surface to volume ratio of the hemorrhage, who have better survival prognostics, may absorb a given blood volume over the larger surface more easily or more rapidly.

The individual predictors derived from the CT image of the patient: surface to volume, volume, sphericity, and surface area may serve as a diagnostic tool for SAH automatic detection.

### 4.2 Diagnosing of SAH

The diagnosis of SAH is currently being done by experienced radiologists within 1 min from opening of the image on the screen. Our method is competitive in the sense that it can be done automatically, *before* the image reaches the radiologist, shortly after it is reconstructed by the CT scanner being sent to a processing unit that processes the image within Docker image. This way, the image may be prioritized in the processing queue for the radiologist.

### 4.3 Vasospasm/DCI

No single predictor of vasospasm/DCI was found significant. To our knowledge, this is the first panel model for SAH that predicts intra-hospital death and vasospasm/DCI from CT image data combined with emergency department clinical evaluation.

Notably, the model predicted correctly the death of five and vasospasm/DCI of two patients, that did not pass thresholds at 80% specificity of individual variables listed in Table 3. These are highly complex image-derived data. They can be initially difficult to understand, especially for the people who do not have daily experience with that type of data.

The comparison of DCI and vasospasm prediction model with available single parameter biomarkers and multivariate models is shown in Table 4. There are only two single biomarkers that give higher AUC (leukocytes count^13^ and serum glucose phosphate ratio^14^ on day of admission), but they have no confidence interval data, and their values lie within our confidence intervals, therefore we can assume that their AUC is not significantly different. All other biomarkers have AUC values that are not significantly different from ours. For other studies there is no data on specificity and sensitivity except for CRIG^10^, but this model has low sensitivity of 44%, compared to our 80% or 90% depending on the threshold and therefore specificity choice. The model of Ramos *et al.*^36^ for DCI prediction is of similar AUC, but it includes data after treatment and operation, so its results are available day(s) after admission. Our model is based on neurological assessment and CT scan data, while CRIG requires blood analysis, results of which come in later stage of the admission.

**Table 4.**
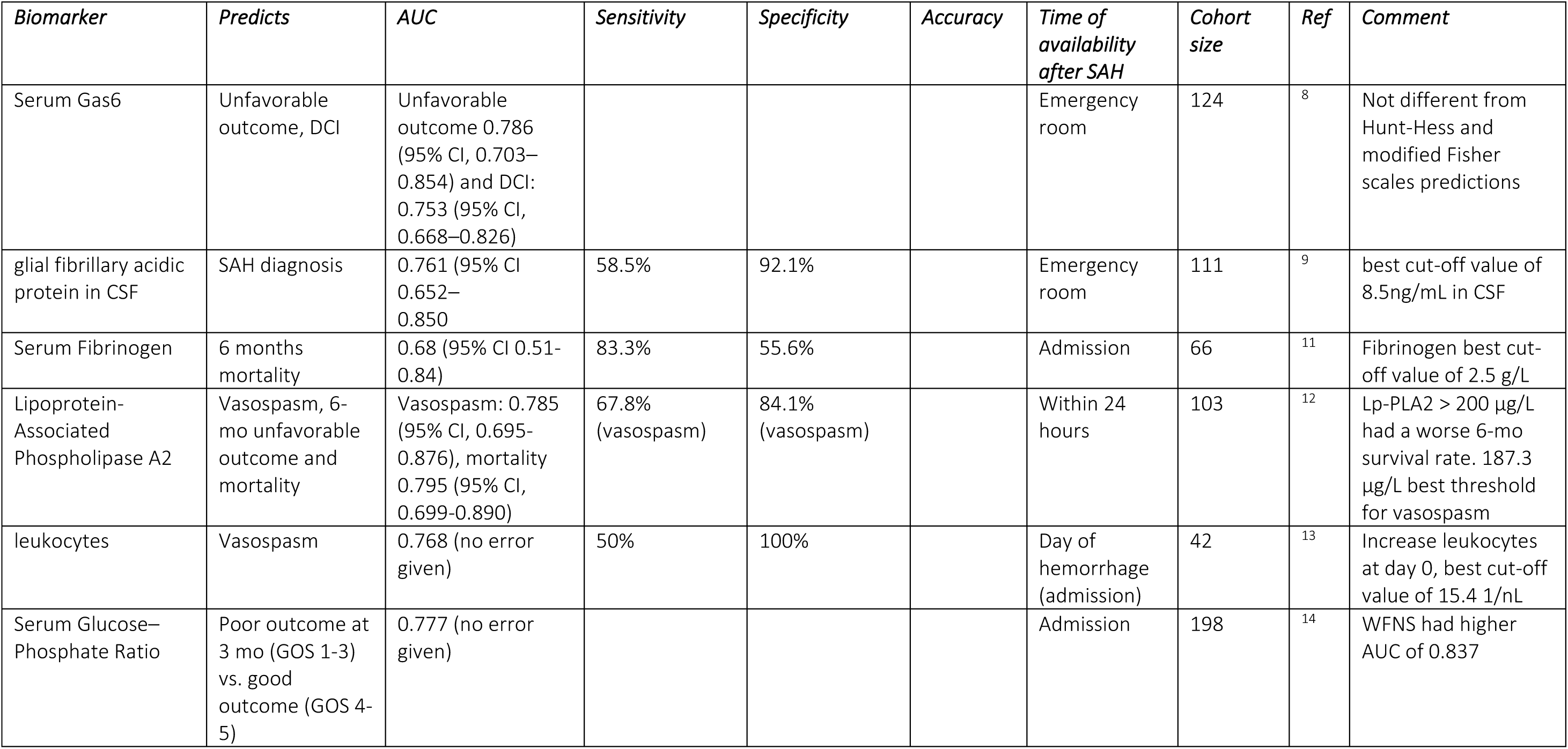
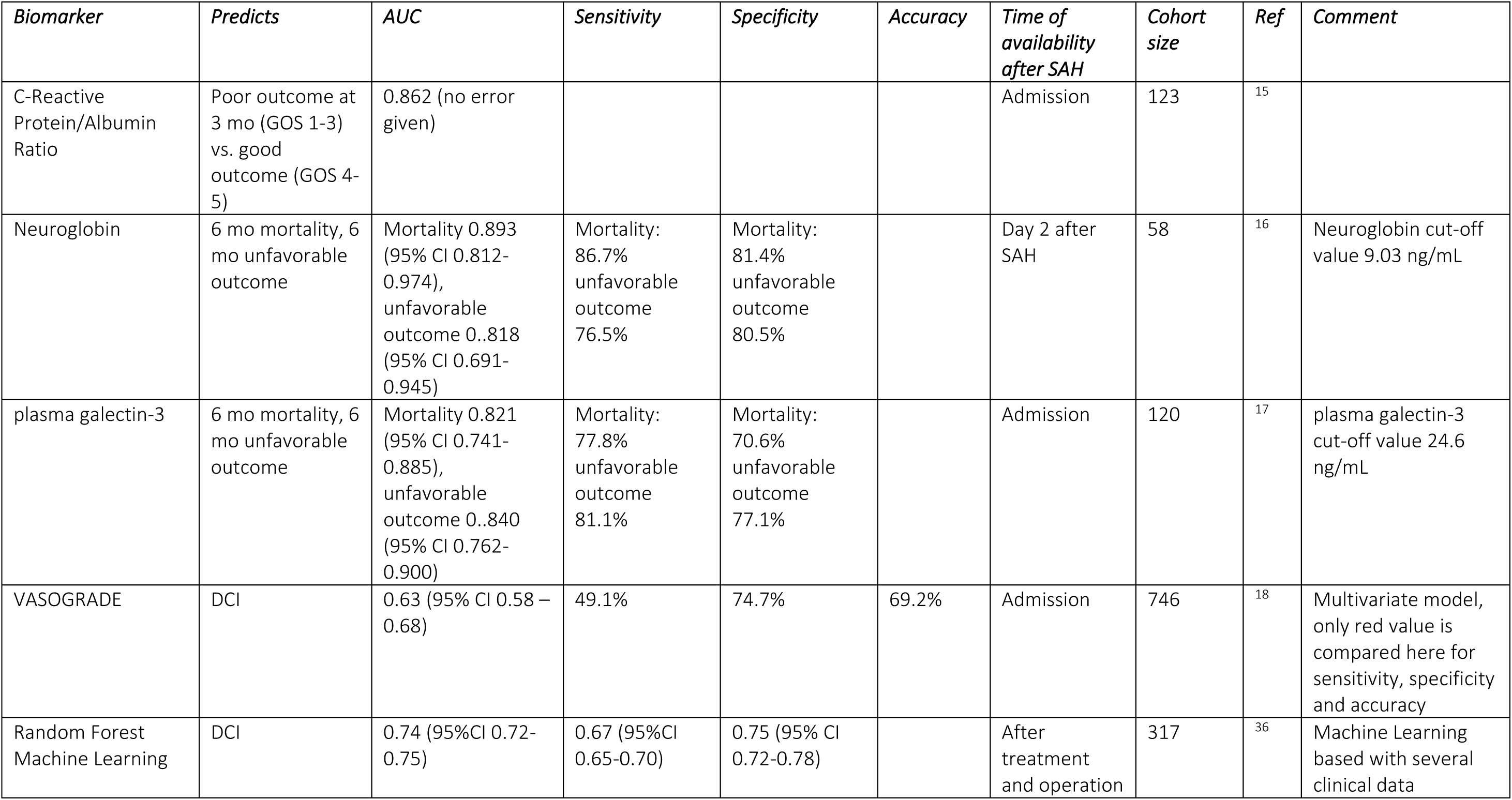
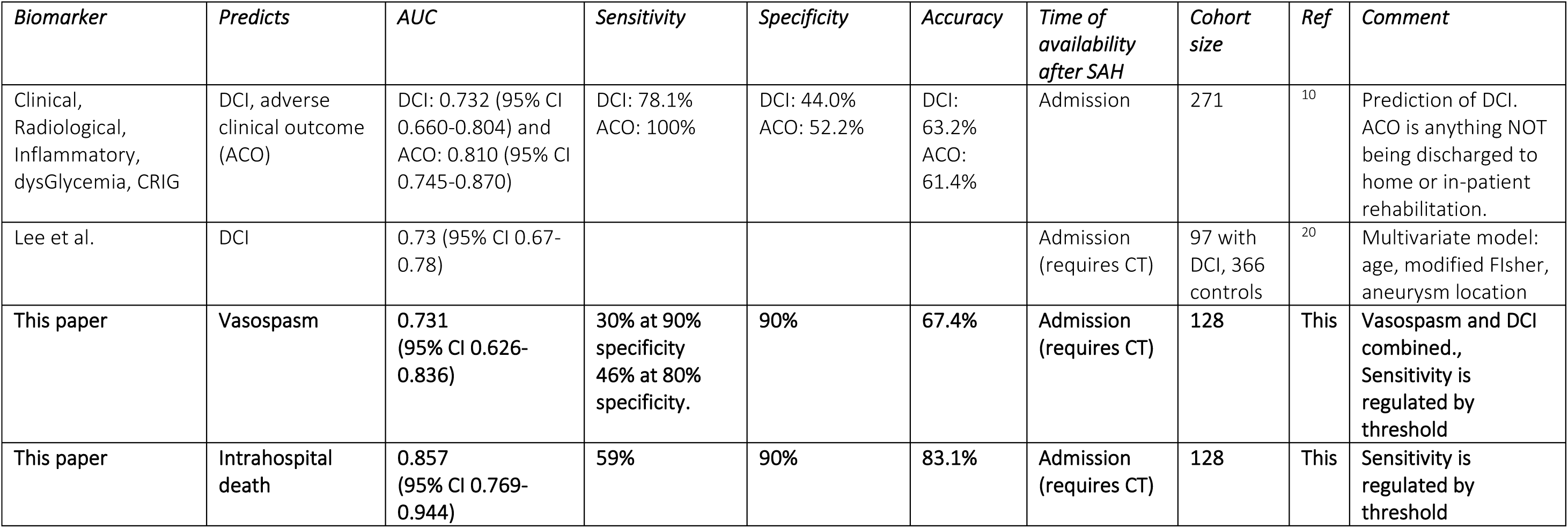
Comparison of predictive biomarkers in SAH with the results of current paper.

### 4.4 Death

Death prediction is improved with the model panel compared with individual predictors. At 90% specificity, the prognostics improved drastically. The common-sense interpretation of this value is that the possible death or survival for *nine* out of ten patients will be predicted correctly with a 59% probability. For a single variable, this probability is only 32% for volume and 36% for surface-to-volume variables. This illustrates the power of the biomarker panel model. In respect of (Nagelkerke) R^2^ value – in mortality prediction, we reached 0.42 compared with the best reported 0.31^23^.

The models developed in this paper perform better in negative predictive value than in positive predictive value. They also perform better for death prediction than for vasospasm. One of the reasons which come up while examining CT images of false negatives for death is that for patients with hemorrhage extending to ventricles, the dilution of the blood in cerebrospinal fluid results in an overall reduction of X-ray density of this spill. Therefore, examining evident hemorrhages in this area, one can see that a denser hemorrhage zone has even 40-50 HU, which is not captured by the current method, where the VOIs were selected at higher HU densities. Extending the hemorrhage definition down to 40 HU results in the selection of healthy areas in the brain, which needs to be avoided.

SAH outcome prediction studies for death, dichotomized Glasgow Outcome Scale (GOS) at 6 months, or Modified Rankin scale (mRS) outcomes were reviewed by Jaja *et al.*^37^ and classified by AUC as performance. The AUC showed there are 0.78-0.86. Our model for death prediction reached 0.857 situating at the upper part of the models.

The prediction of DCI was shown by De Oliveira^4^ with AUC of 0.63 based on WFNS and modified Fisher scales. Another model predicting vasospasm based on blood test one day after onset of SAH is based on Serum high-mobility group box-1 concentration^38^. It reached AUC of 0.704. Our model performed significantly better at AUC of 0.731, and accuracy, and the results are available on the day of SAH.

One single biomarker which is measured for the vasospasm/DCI detection about three days after onset of SAH is S-100B (sometimes referred to as S100β)^39^ ^40^ got high attention as a prognostic of poor outcome and vasospasm/DCI occurrence. However temporal evolution of S-100B is important^39^. S-100B data was available only for a minority of our patients, so they were not used in the analysis.

The automation of the process and the possibility to directly attach it to CT scanner image reconstructing application or making it available in the web app at www.qmenta.com with the evaluation available within a few minutes make it an extremely versatile and powerful tool in the hands of a radiologist or even inexperienced technician.

The results of our model are compared with other models in Table 4. We have chosen an extreme outcome of intrahospital death, while other models chose unfavorable or poor outcome, which usually comprised of GOS values of 1-3 (1-death, 2-persistent vegetative state, 3-severe disability) after 3 or 6 months. Our model concentrates on evaluation of in-patients and therefore hospital management. It is not surprising therefore, that it had the highest AUC, but it lied within confidence intervals of other studies, yet predicting mortality within a much longer period and including survivors with high degree of disability.

Apart from age and sex of the patient surface-to-volume ratio entered the model and was the most significant. An intuitive description of this finding is that for the blood spill of the same volume, the patient that has it with the larger surface can more easily reabsorb the blood and has higher chance of survival. That translates to general surface-to-volume ratio, where even a small volume, but with low surface induces more brain damage and leads to death. Until recently the simple volume was considered as the largest threat, here we indicate that even small spill volume with small surface (i.e. spherical or ellipsoid) is leading to higher death probability.

### 4.5 Limitations

There are some limitations to the current study, including that the sample size is limited, especially the small number of controls. Due to limited number of cases, we pooled together vasospasm and DCI patients together. In this model, there is no count of time between admission and intra-hospital death, nor between admission and vasospasm/DCI. That influences the data on vasospasm/DCI of the patients who died shortly after admission and did not develop vasospasm/DCI. The model does not work properly for CT scans with metal objects within brain area of the skull, that is for some operated patients. Moreover, the CT scans should be of high quality, without visible rays coming from image reconstruction or bone. For the areas close to the gray matter, it should be easily distinguishable from white matter while using −20…80 HU viewing range.

### 4.6 Conclusions

In conclusion, we have developed a model for outcome prediction in SAH in terms of vasospasm/DCI and death. It is based on a CT scan at the emergency department. The unexpected finding that the highest significance of death probability is for surface-to-volume ratio will be studied in the prospective clinical trial. The similar finding for vasospasm/DCI probability for statistical significance of fractal dimension and sphericity will be further studied in the prospective clinical trial.

## Glossary

DCI: Delayed Cerebral Ischemia

SAH: Subarachnoid hemorrhage

WFNS: World Federation Neurological Surgeons

STROBE: STrengthening the Reporting of OBservational studies in Epidemiology

TRIPOD: Transparent Reporting of a multivariable prediction model for Individual Prognosis Or Diagnosis

HH: Hunt-Hess scale

MCA: medial cerebral artery

Vm: average speed

ICA-ext: extracranial internal carotid artery

MNI: Montreal Neurological Institute

HU: Hounsfield Units

NPV: negative predictive value

PPV: positive predictive value

CU: units derived from HU see reference^1^

## Data Availability

All data produced in the present study are available upon reasonable request to the authors.

## Funding

1. The patients for this study were recruited thanks to a grant from Consejería de Igualdad, Salud y Políticas Sociales de Andalucía, Spain (PI-0136-2012).
2. The study was partially funded by the project CI19-00068 from CaixaImpulse program of La Caixa Foundation (Barcelona, Spain). The PI of the project (Marcin Balcerzyk) is the corresponding author of this publication. We acknowledge the valuable comments from Itziar Neve-Lete and Jose Aurelio Cordero-Guevara both from Bioaraba Health Research Institute.

## 8 Supplementary section

### 8.1 Patient details

Patients were admitted to a neurological intensive care unit. They were monitored with daily transcranial Doppler (TCD) ultrasound. The peak systolic velocity, peak diastolic velocity, mean blood flow velocity, pulsatility index, and Lindegaard^41^ ratio were recorded for the middle cerebral, anterior cerebral, posterior cerebral, basilar, and internal carotid arteries. The medial arterial pressure and the pCO2 at the time of the study were recorded. *Vasospasm and DCI were considered* if detected by any of the following modalities: clinically or through TCD and arteriography information. *We defined DCI* as a deterioration of the level of consciousness equal to or exceeding 2 points of the GCS or the appearance of the focal deficit, between the 4^th^ and the 14^th^ day of the hemorrhage, not related to rebleeding, hematoma, hydrocephalus, or metabolic alterations^6^, The information was taken from the medical records. *TCD vasospasm* of the medial cerebral artery (MCA) was defined as at the existence of mean velocity (Vm,) level MCA exceeding 120 cm/s with a hemispheric index value equal to or greater than 3, starting from the 4^th^ day after bleeding. A hemispheric index Vm_MCA_/Vm_ICA-ext_ is a ratio between MCA Vm (Vm_MCA_) and the Vm of the extracranial internal carotid artery, (ICA-ext) homolateral at the submaxillary level (Vm_ICA-ext_)^41^. Angiographic vasospasm was diagnosed by catheter arteriography and radiology interpretation. Patients with angiographic narrowing of at least one artery were considered to have vasospasm. The arteriography was done depending on clinical examination/TCD results. All sets of vasospasm/DCI patients were pooled together due to the low number of patients for model training. Vasospasm (VSP) refers to the radiologic or TCD phenomenon of narrowing of the brain arteries and DCI is neurological deterioration 3 to 14 days after SAH not due to identifiable causes like seizure, infection, hyponatremia, etc. VSP is common after SAH; about 67% of patients get it but not all of them get DCI (DCI only happens in about 33% of patients). So, there can be a VSP without DCI. VSP must be quite severe and usually involve multiple arteries before a patient will get DCI from it. About 50% of patients with severe VSP get DCI, and very few with mild or moderate VSP get DCI. Also, rarely (about 5% of DCI cases) DCI happens in a patient with no or mild VSP^42^.

Management of patients included repair of the aneurysm by endovascular coiling or surgical clipping within 72 hours of admission. All patients received oral nimodipine. (a calcium antagonists). If DCI was suspected, then induced hypertension, hemodilution, and hypervolemia were used. When there was an inadequate clinical response to hemodynamic therapy, mechanical, pharmacological or endovascular techniques were implemented.

**Suppl. Figure 1.**
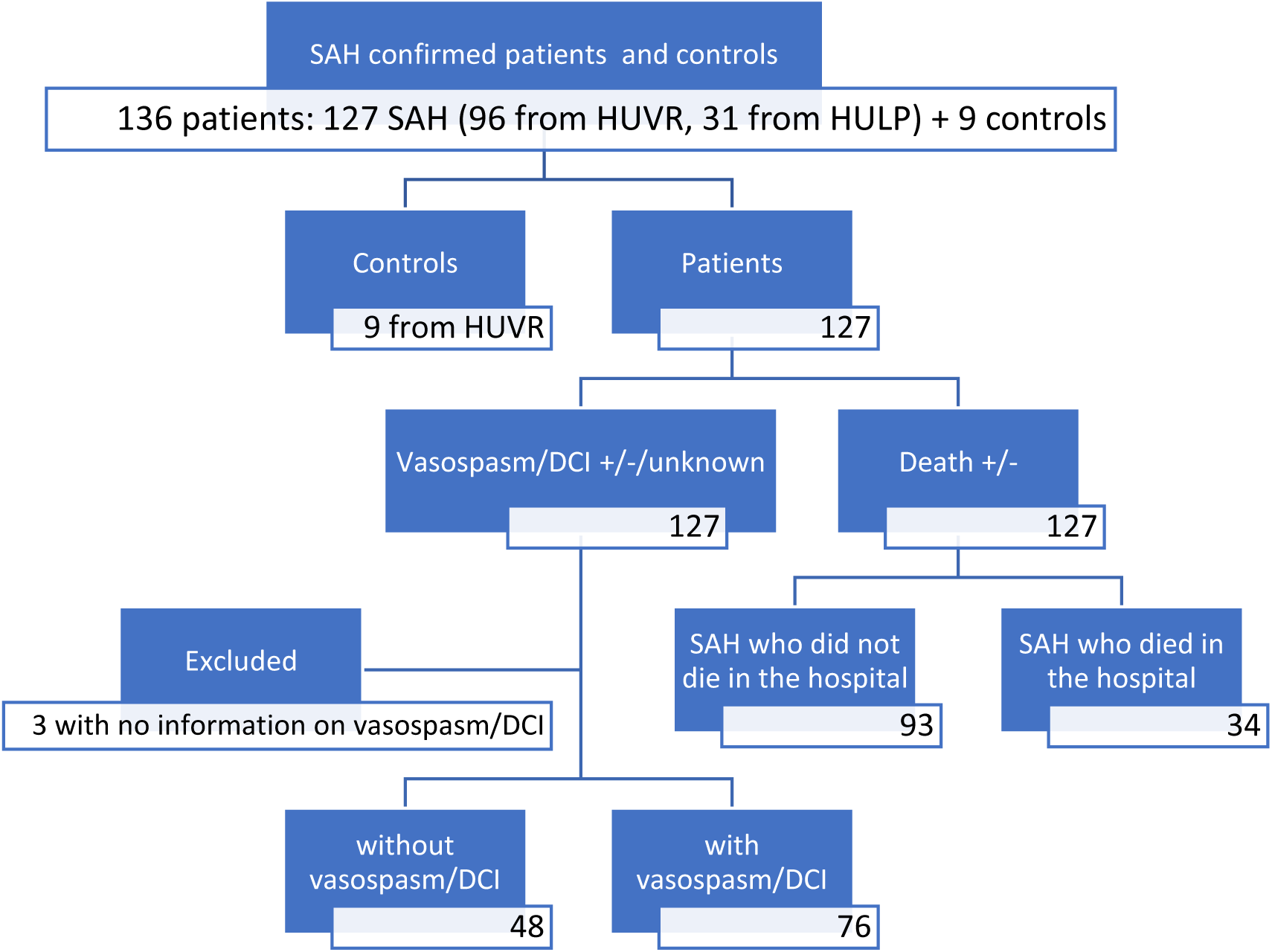
Study flow chart of patients and controls. For training 70% randomly selected patients were used, the remaining 30% were used for validation.

### 8.2 Standard Protocol Approvals, Registrations, and Patient Consents

This research project was overseen and approved by Hospital Universitario Virgen del Rocio in Seville, Spain hospitaĺs ethics committee (Cod. CEI2012PI/228). Informed consent was obtained from all patients participating in the study or from their close relatives. The corresponding author declares taking full responsibility for the data, the analyses, and interpretation, and the conduct of the research; that he has full access to all the data; and that he has the right to publish any and all the data, separate and apart from the guidance of any sponsor.

### 8.3 Image processing details

The processing of CT images in PMOD software is the following:

1. The image was loaded and normalized to Montreal Neurological Institute (MNI) based CT template of 1 mm uniform resolution^24^. The image deformation was plastic, but this way permitted the comparison between subjects of different skull and brain sizes. Notably, the Hounsfield Units (HU) were converted to CU units^1^, enhancing the contrast of the brain between −99 and 100 HU. It is crucial for correct registration to the CT template. The resulting image was converted back to HU units.
2. A brain mask with smooth transition edges with values 0 outside and 1 inside of the brain was used at 0.85 value to delineate the brain, and within this region, the segmentation in three segments was done: 60-80 HU, 50-60 HU, 80-85 HU, resulting in three Volumes of Interest (VOIs) per image. The values 60-80 HU best correspond to hemorrhage in CT images. The VOIs were calculated purely based on the value, so there were some areas with isolated pixels, especially close to the skull.
3. The three segmented VOIs were analyzed for the following parameters, then used as predictors:

a. Volume (VOL, mL), for the pixels selected to be included in VOI, their volumes were summed up.
b. Surface area (SRA, cm^2^), the surface of each pixel that was included in the VOI was checked if its surface neighbors to the other pixel in VOI. If the surface pointed to the pixel not included in the VOI, the surface was added to the total surface.
c. Sphericity (SPH, shortly, how in a scale 0 to 1 the object is like a sphere, SPH). The definition of sphericity is 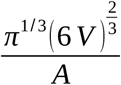, where V is object volume, and A is its surface. For the reasons of the calculation precision, temporarily Sphericity was multiplied by 100 and therefore given in %, and the Surface Area was divided by 1000. Only VOI of 60-80 HU was used.
d. The maximum diameter of the VOI, that is the maximum distance between the VOI pixels (DMX, cm). For each pair of pixels of the VOI, the distance was checked, and the maximum was found.
e. The fractal dimension of the VOI object (FDR). Briefly, the dimension of the point is zero of a string is one of a paper sheet, is two and of the ball is three. Some objects seem to occupy less space than their outer extent does. The examples are a snowflake on a plane, cauliflower, upper branches of the tree, or its root in 3D space. Snowflake fractal dimension is a value between one and two, the dimension of cauliflower is between two and three. For the PMOD fractal dimension calculation, box-counting method^9^ was used.
f. Surface to volume ratio (SRAVOL), which was calculated from points 3.b and 3.a in cm^-1^ units. In chemistry, this parameter is important in reactions. That is used in catalysis using porous materials^43^. Many reactions take place on the surface of the vessel or catalyst. In small volumes, when the reaction takes place on the surface of the vial, reactions normally occur faster. In our case, the diffusion through the surface of the hemorrhage may give importance to this variable.

### 8.4 Statistical analysis details

We constructed several models of biomarker panels (image derived values together with clinical ones, see Supplementary Section 8.6 for the list) for vasospasm/DCI and in-hospital-death using binary logistic regression with forward and backward conditional methods. This modeling was prepared to improve AUC/ROC values for single independent variables.

Predictors were selected for subsequent iteration if p≤0.1 and removed if p>0.1. This way for backward regression, we could eliminate biomarkers which added marginal descriptive value. For forward regression starting from the one with the highest significance, we reach the set, where adding a new biomarker did not significantly change the model. All image derived PMOD predictors and age were treated as continuous. Hunt-Hess, WFNS, and Fisher were treated as ordinal variables and sex as a nominal variable.

The primary minimum limit for the sample size was at least ten cases for each of the individual predictors in a final converged model. At the worst case of the model with 4 parameters (vasospasm probability), we had 21 cases per parameter in a final model. We validated internally with a bootstrapping for a realistic estimation of the performance of all prediction models (i.e. 1: death; 2 and 3: vasospasm) in similar future patients. We repeated the entire modeling process including variable selection in all logistic regression in 100, 200, and 1000 samples were drawn with replacement from the original sample. We determined the performances of the selected prediction model and the simple rules that were developed from each bootstrap in the original sample. Performance measures included the average area under the AUC/ROC (Area under curve/receiver operating characteristics) curve, sensitivity, and specificity for both outcome measures (i.e. vasospasm/DCI and death). We validated this by using both SPSS and STATA. Additionally, a predictive test was carried out using a cross-validation process using holdout samples. This evaluation allows to know whether the model can generate accurate predictions of new interpretable observations. We chose from the sample 75 cases for training, and 10 cases for the test with 10 times of 10 random repetitions. We validated the results by dividing randomly the group of 127 patients in 70%/30% - model training/validation groups. ROC curves for training and validation groups were tested for the statistical difference with the bootstrap test in the pROC package.

For the developed biomarker panel model, in the case of Backward regression, the full set of variables for all three VOI HU ranges and all clinical variables was selected at the beginning of the iteration. The entry probability for entering the panel was 0.05 and removal probability was 0.1. Constant was included in the model. We follow a typical definition of accuracy as a percentage fraction of the correct prediction of the total group. Statistical significance was assumed at p<0.05. Confidence intervals (CI) were set at 95%.

### 8.5 Example patient data

Below we showed four examples of four types of patients with a different clinical outcome of survival and vasospasm/DCI. The typical shape of the VOI for 60-80 HU of the patient ([36…40]-year-old male) with vasospasm/DCI who died in the hospital is shown in 3D rendering in yellow in Suppl. Figure 2 (a). This patient had a hemorrhage which measured 23.03 mL volume in the 60-80 HU VOI range. One can see that the VOI is very fragmented, that it is located primarily in the space between hemispheres, that resulted in a surface area of 777 cm^2^, low sphericity of 5.03%, diameter which is close to the brain diameter (17.58 cm), the surface to volume ratio of 33.75 cm^-1^ and fractal dimension of 1.88. The model prediction for Death is 0.550>0.5 so it predicts death (true positive), and 0.614>0.5 it predicts vasospasm/DCI occurrence (true positive). The patient had a Hunt-Hess score of 5 and WFNS of 5. The patient did not meet any individual image derived predictor for Death characteristics, only Hunt-Hess, and WFNS.

The 3D rendering of a patient ([46…50]-year-old female) with vasospasm/DCI who did not die in the hospital is shown in Suppl. Figure 2 (b). This patient had a hemorrhage which measured 22.29 mL in the 60-80 HU VOI range, had vasospasm/DCI, and did not die in the hospital. One can see that the VOI is fragmented, which resulted in a surface area of 717 cm^2^, low sphericity of 5.3%, diameter which is close to the brain diameter (17.26 cm), the surface to volume ratio of 32.17 cm^-1^ and fractal dimension of 1.94. The model prediction for Death is 0.151<0.5 so it predicts survival (true negative), and with 0.666>0.5 it predicts vasospasm/DCI occurrence (true positive). The patient had a Hunt-Hess score of 5 and WFNS of 4. He did not meet any individual imaged derived predictors for death neither at 80% nor at 90% specificity, met both Hunt-Hess and WFNS for both death and of vasospasm/DCI.

The 3D rendering of a patient ([66…70]-year-old male) without vasospasm/DCI who died in the hospital is shown in Suppl. Figure 2 (c). This patient had a hemorrhage which measured 95.01 mL in the 60-80 HU VOI range, did not have vasospasm/DCI, and died in the hospital. One can see that the VOI has a voluminous but discontinuous part in the medial part. VOI has a surface area of 1658 cm^2^, the sphericity of 6.07%, diameter which is close to the brain diameter (17.71 cm), the surface to volume ratio of 17.44 cm^-1^ and fractal dimension of 2.11. The model prediction for Death is 0.762>0.5 so it predicts death (true positive), and 0.041<0.5 it predicts no vasospasm/DCI occurrence (true negative). Notably, the patient had a Hunt-Hess score of 1 and WFNS of 2. From individual Death predictors, the patient passed the threshold for 90% specificity in the surface to volume ratio, volume, fractal dimension, and with 80% specificity the threshold of sphericity.

The 3D rendering of a patient without vasospasm/DCI who survived in the hospital is shown in Suppl. Figure 2 (d). The patient was diagnosed with SAH. The patient ([66…70]-year-old male) had a zone of 60-80 HU with the volume of 18 mL, the surface of 851 cm^2^, low sphericity of 3.9%, a diameter which is close to the brain cm diameter (17.4 cm), the ratio of surface to volume of 46.4 cm^-1^ and fractal dimension of 1.61. The prediction of the model for the death at the hospital gave 0.234 < 0.5 what predicts survival (true negative) and 0.197 < 0.5 predicts the non-occurrence of vasospasm/DCI (true negative). The patient had Hunt-Hess 2 and the WFNS 2 score. The patient did not meet any individual predictor derived from the image of the characteristics of death in 80% of specificity.

**Suppl. Figure 2.**
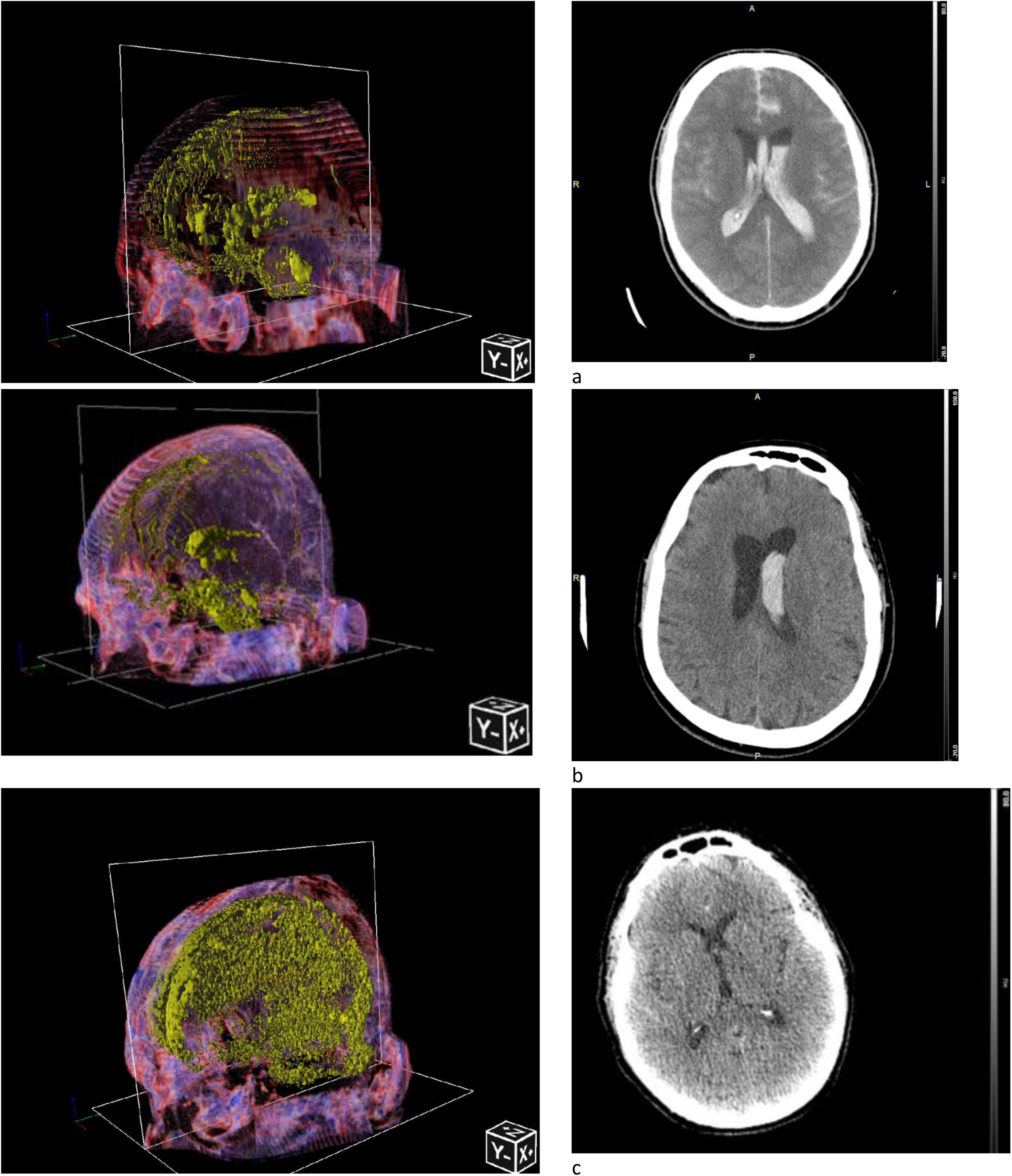
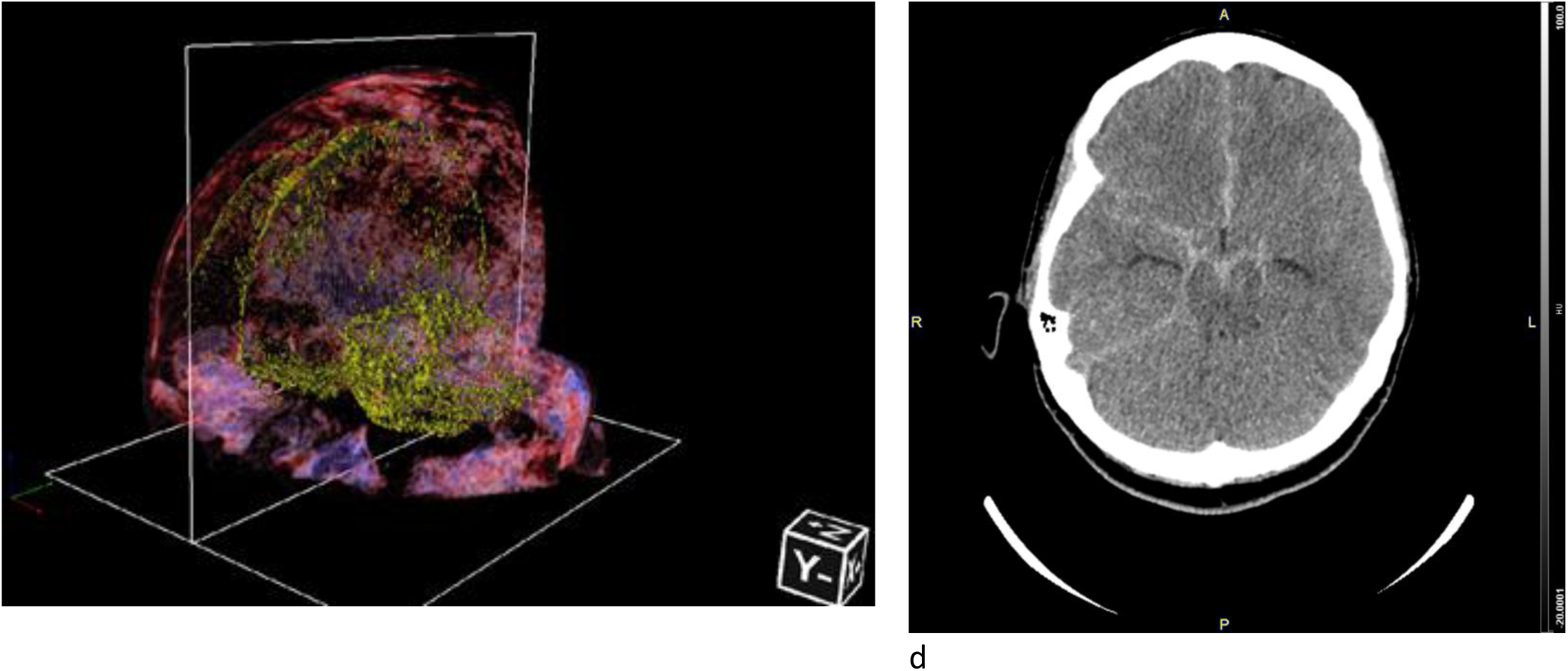
Examples of four types of patients with a different clinical outcome of survival and vasospasm/DCI. On the left, the 3D rendering of normalized is shown. On the right, the transversal section is shown from the original CT at the most pronounced zone of SAH. (a) Patient with VOI volume of 23.03 mL (yellow), who died in the hospital and had vasospasm/DCI. Z+ is head direction, X+ is left of the patient, Y-is the anterior direction of the patient. (b) Patient with VOI volume of 22.29 mL (yellow), who did not die in the hospital and had vasospasm/DCI. (c) Patient with VOI volume of 95.05 mL (yellow), who died in the hospital and did not have vasospasm/DCI. A high number of yellow voxels close to the skull is a confirmed SAH (d) Patient with a VOI volume of 19.13 mL (yellow) who survived in the hospital and did not have vasospasm/DCI but was diagnosed with SAH. There are supplementary video files for the 3D rendering of each patient.

### 8.6 Tables of AUC/ROC and model parameters

**Suppl. Table 1.**
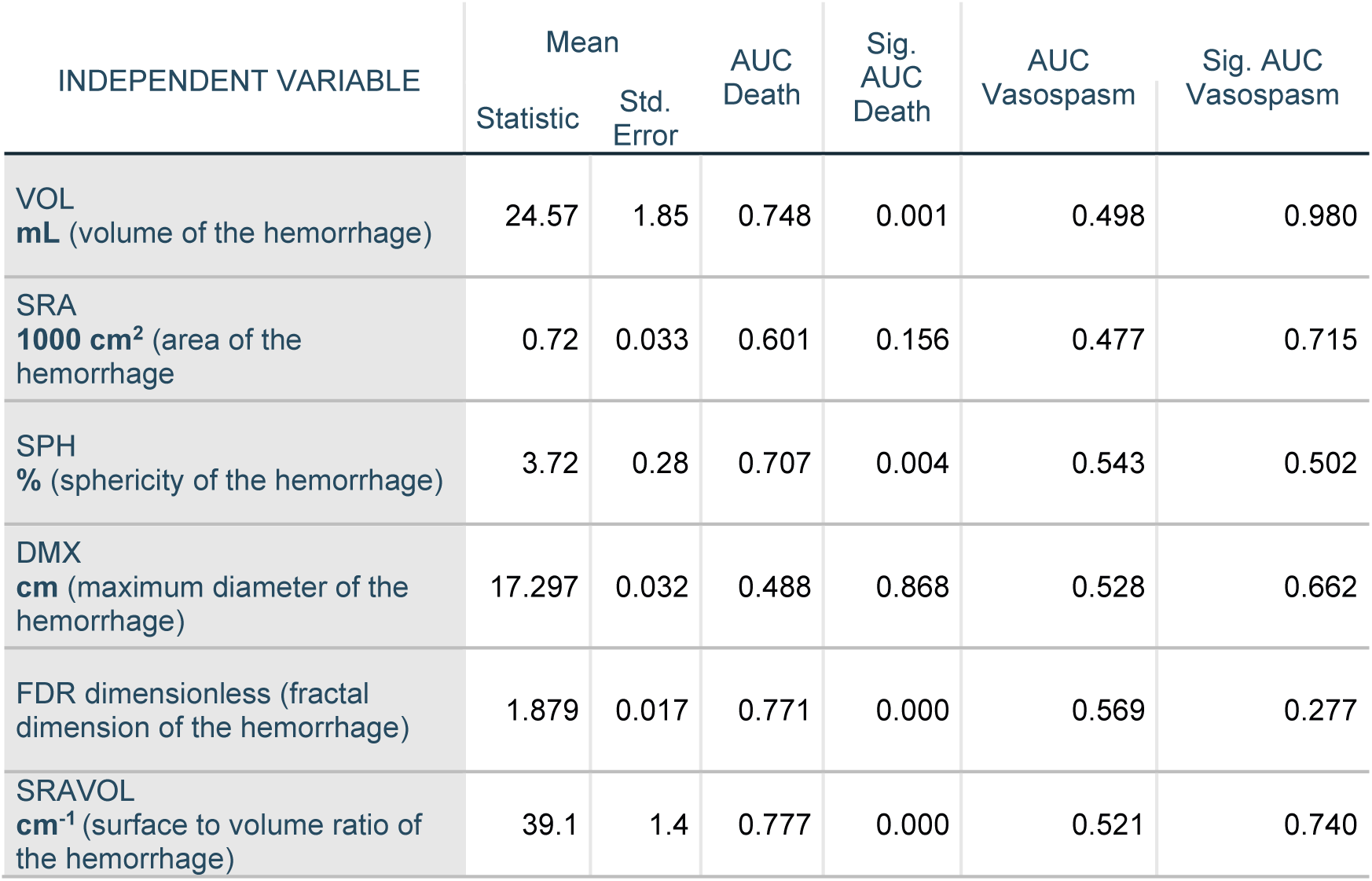
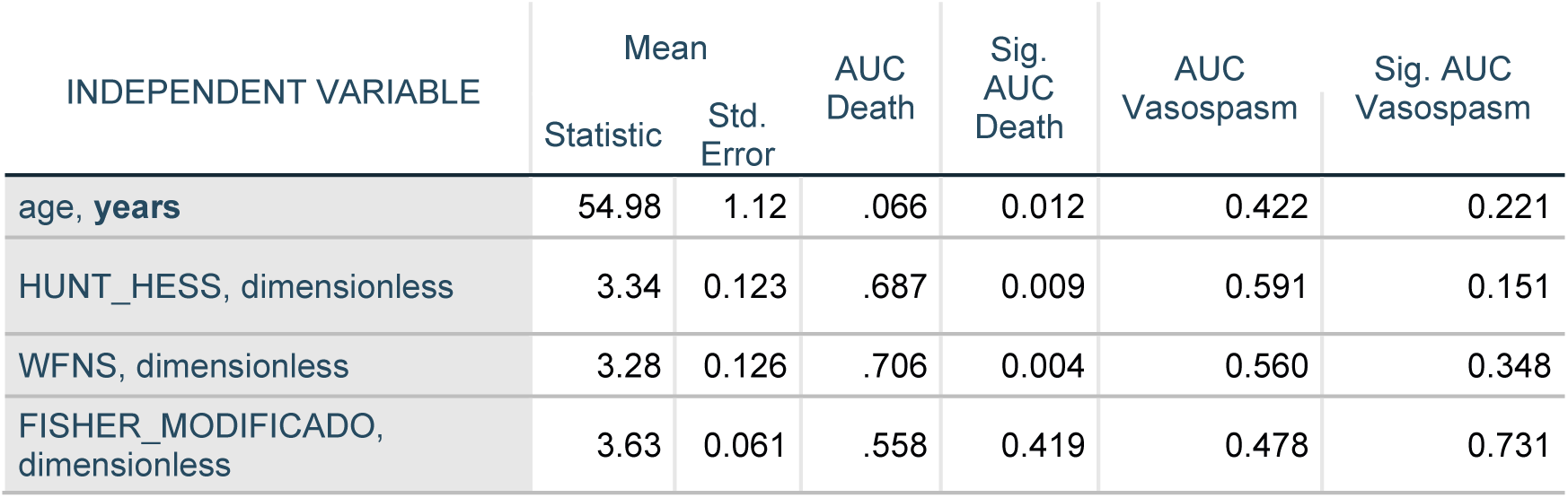
Descriptive statistics of image-derived and clinical variables and AUC/ROC analysis for Death and Vasospasm.

| INDEPENDENT VARIABLE | Mean |  | AUC Death | Sig. AUC Death | AUC Vasospasm | Sig. AUC Vasospasm |
| --- | --- | --- | --- | --- | --- | --- |
|  | Statistic | Std. Error |  |  |  |  |
| VOL<br>mL (volume of the hemorrhage) | 24.57 | 1.85 | 0.748 | 0.001 | 0.498 | 0.980 |
| SRA<br>1000 cm<sup>2</sup> (area of the hemorrhage) | 0.72 | 0.033 | 0.601 | 0.156 | 0.477 | 0.715 |
| SPH<br>% (sphericity of the hemorrhage) | 3.72 | 0.28 | 0.707 | 0.004 | 0.543 | 0.502 |
| DMX<br>cm (maximum diameter of the hemorrhage) | 17.297 | 0.032 | 0.488 | 0.868 | 0.528 | 0.662 |
| FDR dimensionless (fractal dimension of the hemorrhage) | 1.879 | 0.017 | 0.771 | 0.000 | 0.569 | 0.277 |
| SRAVOL<br>cm<sup>-1</sup> (surface to volume ratio of the hemorrhage) | 39.1 | 1.4 | 0.777 | 0.000 | 0.521 | 0.740 |

| INDEPENDENT VARIABLE | Mean |  | AUC<br>Death | Sig.<br>AUC<br>Death | AUC<br>Vasospasm | Sig. AUC<br>Vasospasm |
| --- | --- | --- | --- | --- | --- | --- |
|  | Statistic | Std.<br>Error |  |  |  |  |
| age, **years** | 54.98 | 1.12 | .066 | 0.012 | 0.422 | 0.221 |
| HUNT_HESS, dimensionless | 3.34 | 0.123 | .687 | 0.009 | 0.591 | 0.151 |
| WFNS, dimensionless | 3.28 | 0.126 | .706 | 0.004 | 0.560 | 0.348 |
| FISHER_MODIFICADO,<br>dimensionless | 3.63 | 0.061 | .558 | 0.419 | 0.478 | 0.731 |

**Suppl. Table 2.**
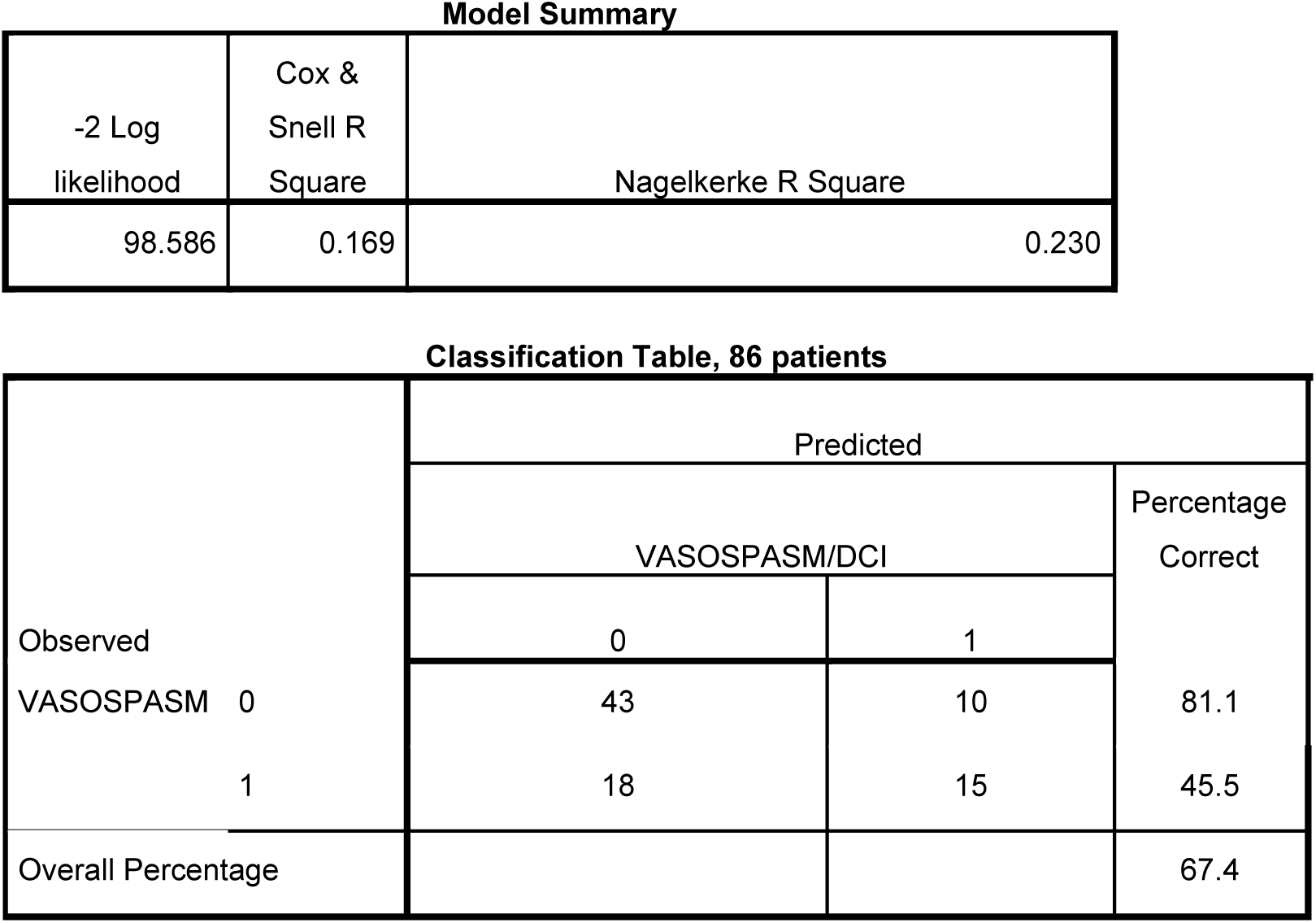
Vasospasm/DCI backward model final step characteristics.

**Suppl. Table 3.**
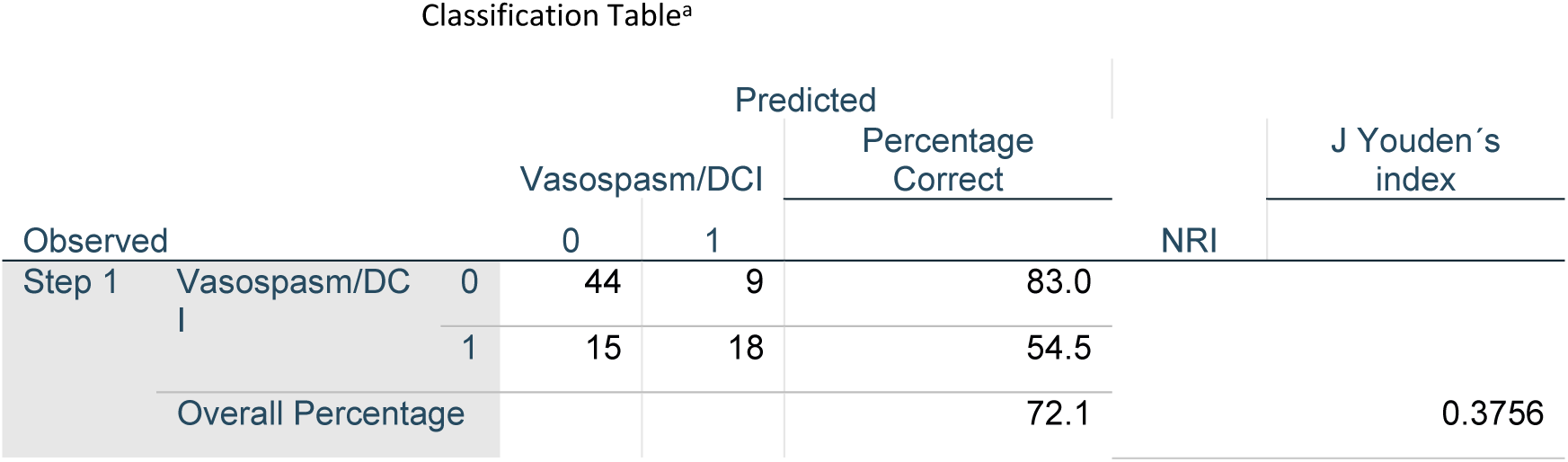
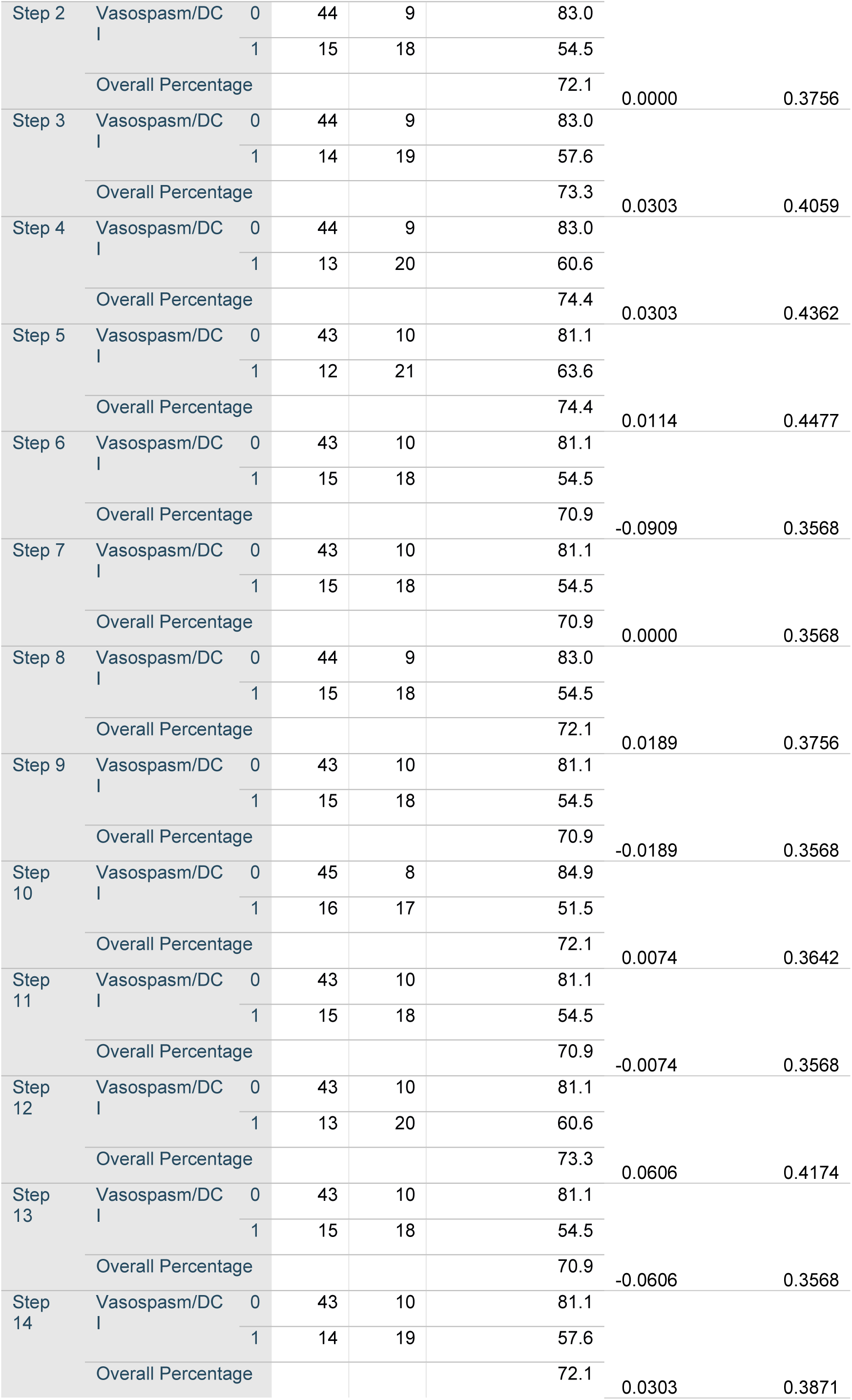
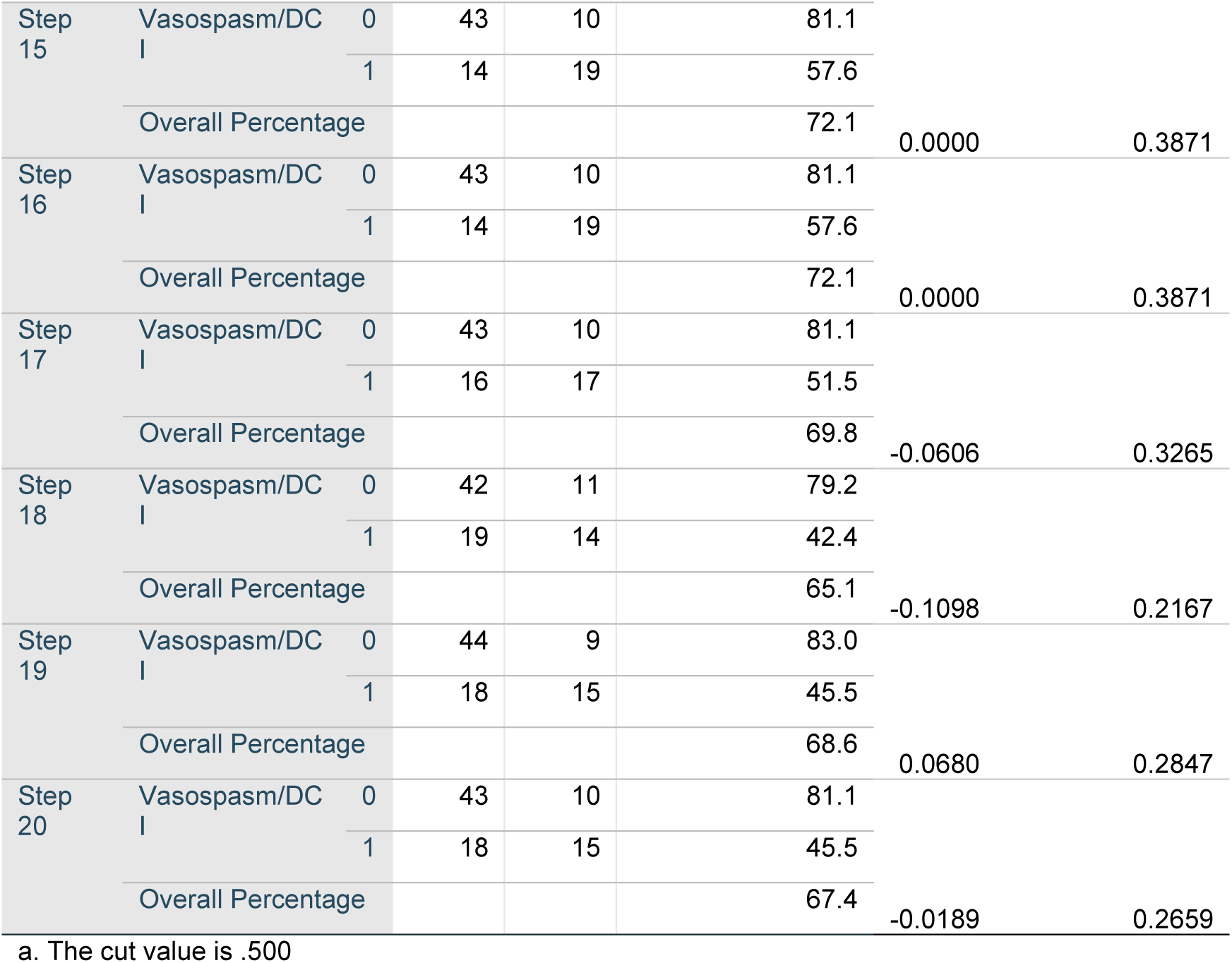
Vasospasm/DCI backward model characteristics – step by step change. Net Reclassification index – NRI is calculated as the difference between step 2 and step 1, etc. in Youdeńs J index.

**Suppl. Table 4.**
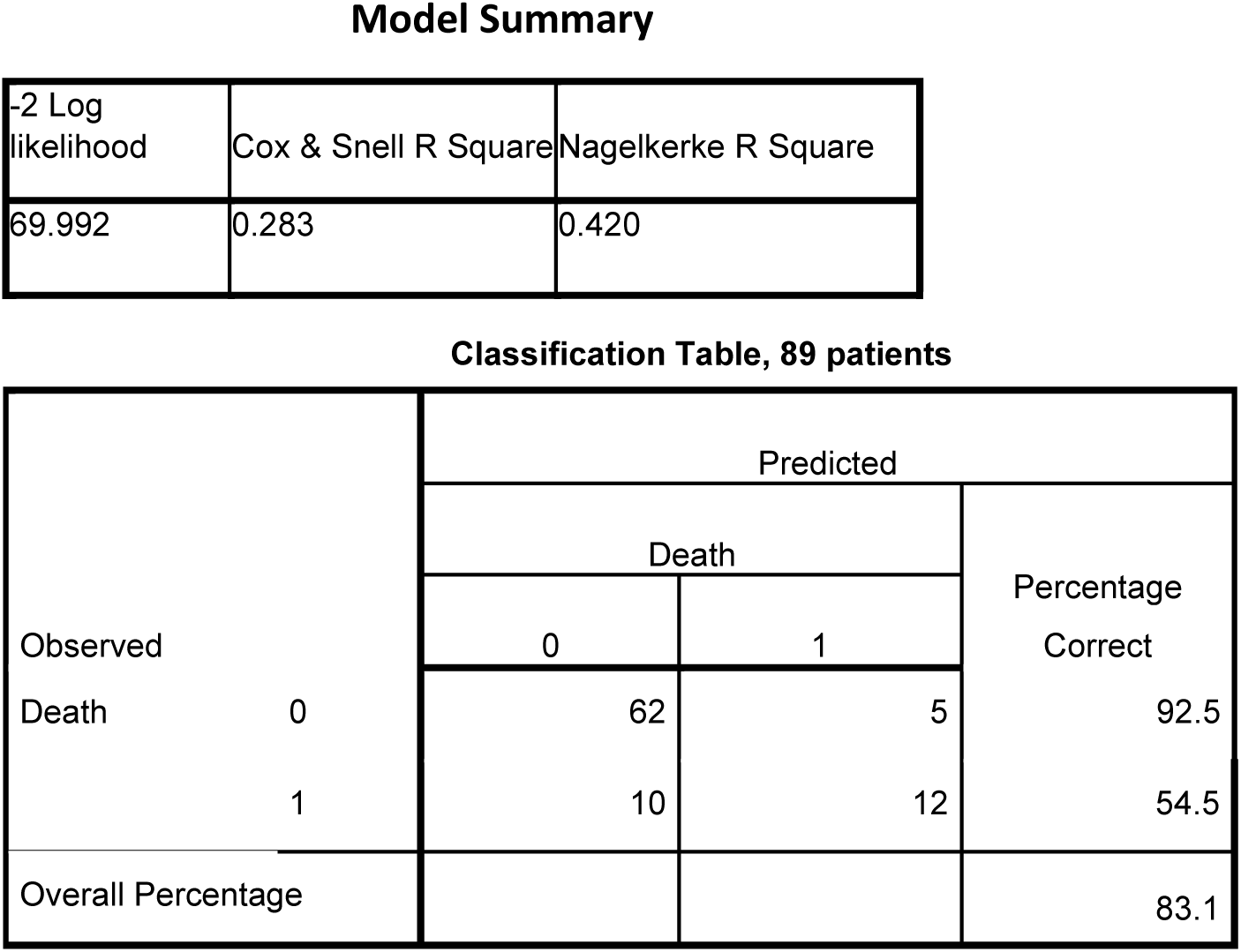
Death in the hospital model final step characteristics.

**Suppl. Table 5.** Death forward model characteristics – step by step change. Net Reclassification index – NRI is calculated as the difference between step 2 and step 1, etc.

Classification Table<sup>a</sup>
| Observed |  |  | Predicted |  | Percentage Correct | NRI | J Youden's index |
| --- | --- | --- | --- | --- | --- | --- | --- |
|  |  |  | Death_Hospital<br>0 | 1 |  |  |  |
| Step 1 | Death_Hospital | 0 | 62 | 5 | 92.5 |  | 0.1981 |
|  |  | 1 | 16 | 6 | 27.3 |  |  |
|  | Overall Percentage |  |  |  | 76.4 |  |  |
| Step 2 | Death_Hospital | 0 | 63 | 4 | 94.0 | 0.1058 | 0.3039 |
|  |  | 1 | 14 | 8 | 36.4 |  |  |
|  | Overall Percentage |  |  |  | 79.8 |  |  |
| Step 3 | Death_Hospital | 0 | 62 | 5 | 92.5 | 0.1669 | 0.4708 |
|  |  | 1 | 10 | 12 | 54.5 |  |  |
|  | Overall Percentage |  |  |  | 83.1 |  |  |

a. The cut value is .500

**Suppl. Table 6.** The accuracy of prediction for validation group with bivariate panel model. Accuracy of Vasospasm model is 57% of Death model is 73% for the validation set.

| Vasospasm/DCI | Predicted No | Predicted Yes | Percent correct |
| --- | --- | --- | --- |
| Observed No | 19 | 4 | 82.6% |
| Observed Yes | 12 | 3 | 20.0% |
| Death | Predicted No | Predicted Yes | Percent correct |
| Observed No | 23 | 3 | 88.5% |
| Observed Yes | 7 | 5 | 41.7% |

**Suppl. Table 7.** The comparison of SAH control, train and validation groups.

| Variables | Control<br>N = 9 | Train<br>N = 89 | Valid<br>N = 38 | p-<br>value<sup>1</sup> | Control<br>vs. Train | Control<br>vs. Valid | Train<br>vs.<br>Valid |
| --- | --- | --- | --- | --- | --- | --- | --- |
| **Volume** |  |  |  | <0.001 | 0.021 | 0.003 | 0.10 |
| Mean<br>(SD) | 6.32<br>(3.35) | 22.46<br>(17.66) | 28.85<br>(25.76) |  |  |  |  |
| Median<br>(Q1, Q3) | 5.68<br>(3.58,<br>8.98) | 17.60<br>(11.60,<br>25.78) | 19.43<br>(11.73,<br>34.90) |  |  |  |  |
| **Surface area** |  |  |  | 0.016 | 0.022 | 0.003 | 0.092 |
| Mean (SD) | 461.57<br>(215.44) | 679.40<br>(238.24) | 768.21<br>(340.94) |  |  |  |  |
| Median (Q1, Q3) | 443.73<br>(300.34, 612.05) | 679.61<br>(546.29, 770.03) | 726.22<br>(543.75, 896.14) |  |  |  |  |
| **Surface to volume** |  |  |  | <0.001 | <0.001 | <0.001 | 0.3 |
| Mean (SD) | 75.68<br>(8.43) | 39.57<br>(15.17) | 36.81<br>(15.60) |  |  |  |  |
| Median (Q1, Q3) | 73.78<br>(73.23, 78.07) | 39.12<br>(29.18, 52.56) | 36.45<br>(25.26, 47.05) |  |  |  |  |
| **Sphericity** |  |  |  | <0.001 | 0.010 | 0.012 | 0.9 |
| Mean (SD) | 0.04<br>(0.01) | 0.05<br>(0.02) | 0.05<br>(0.02) |  |  |  |  |
| Median (Q1, Q3) | 0.04<br>(0.03, 0.04) | 0.05<br>(0.04, 0.06) | 0.05<br>(0.04, 0.06) |  |  |  |  |

<sup>1</sup>Kruskal-Wallis rank sum test

## Appendix 1: Authors

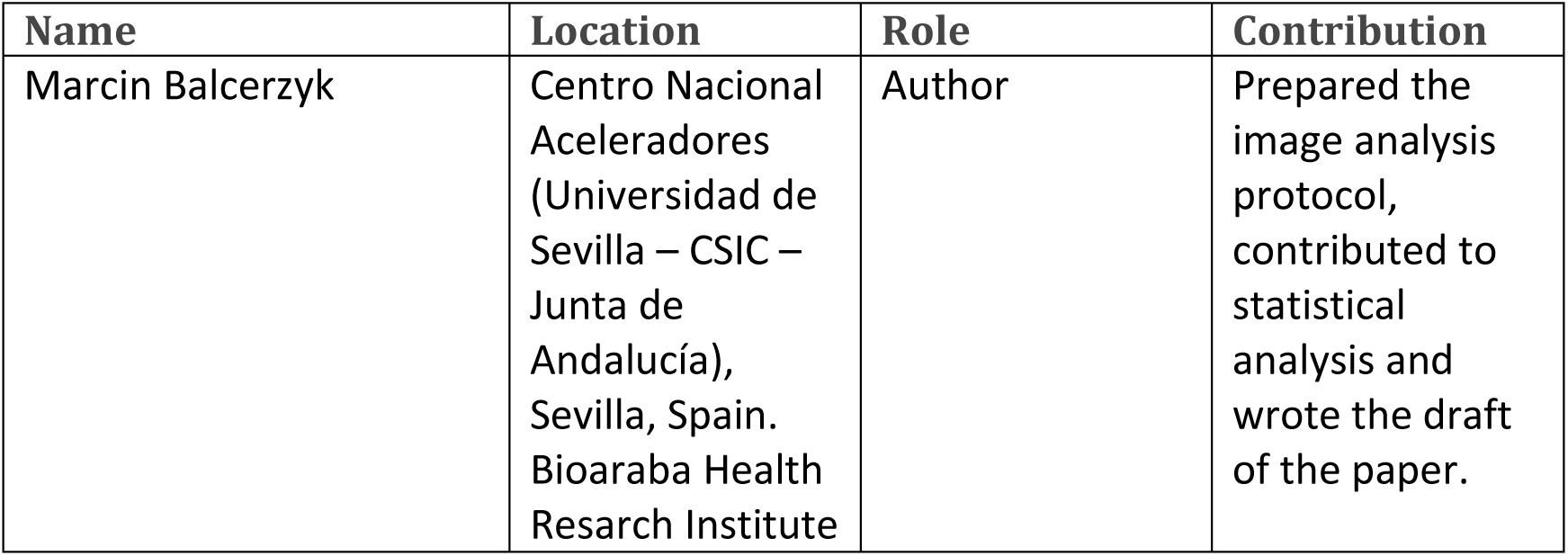
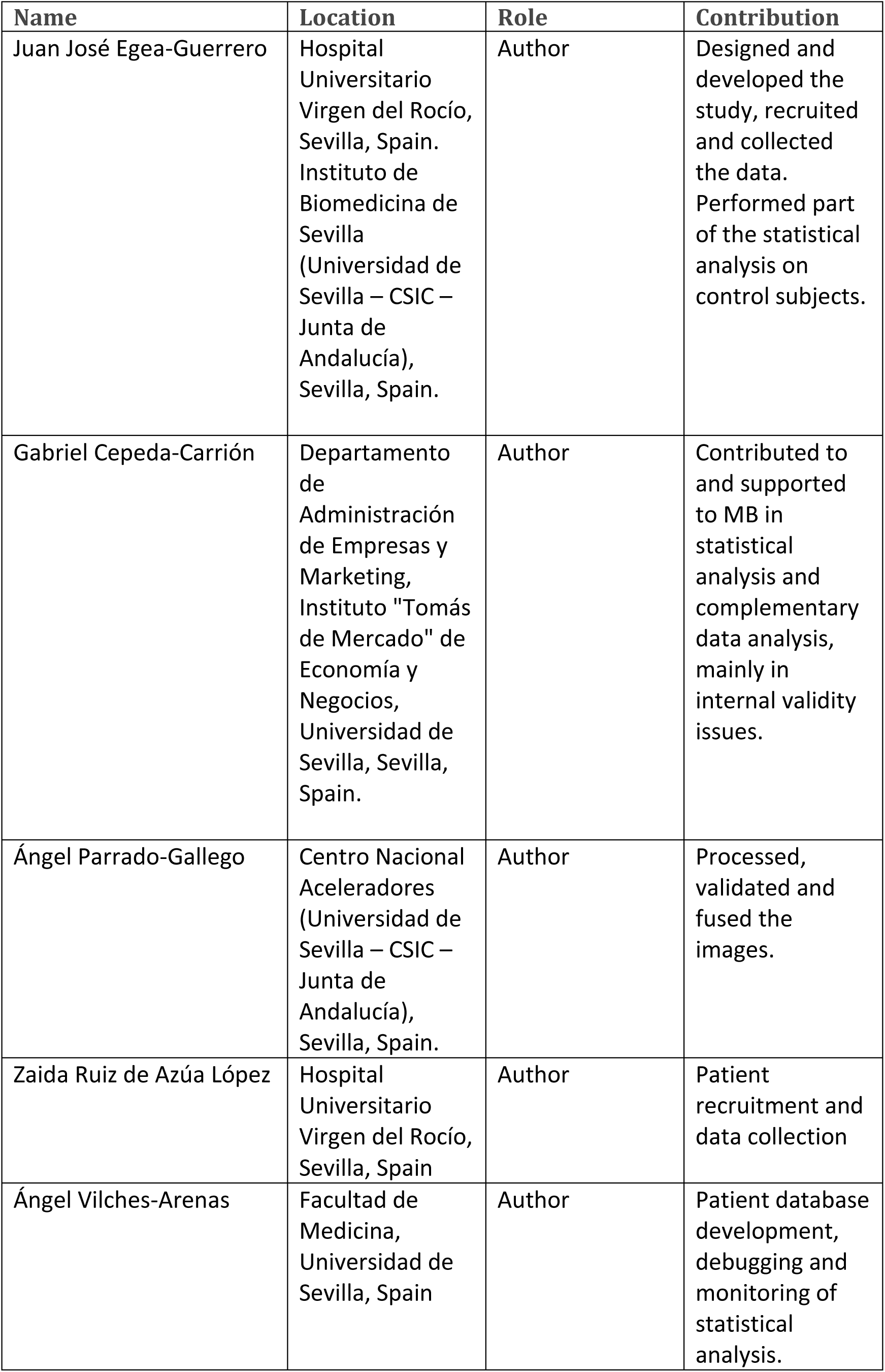
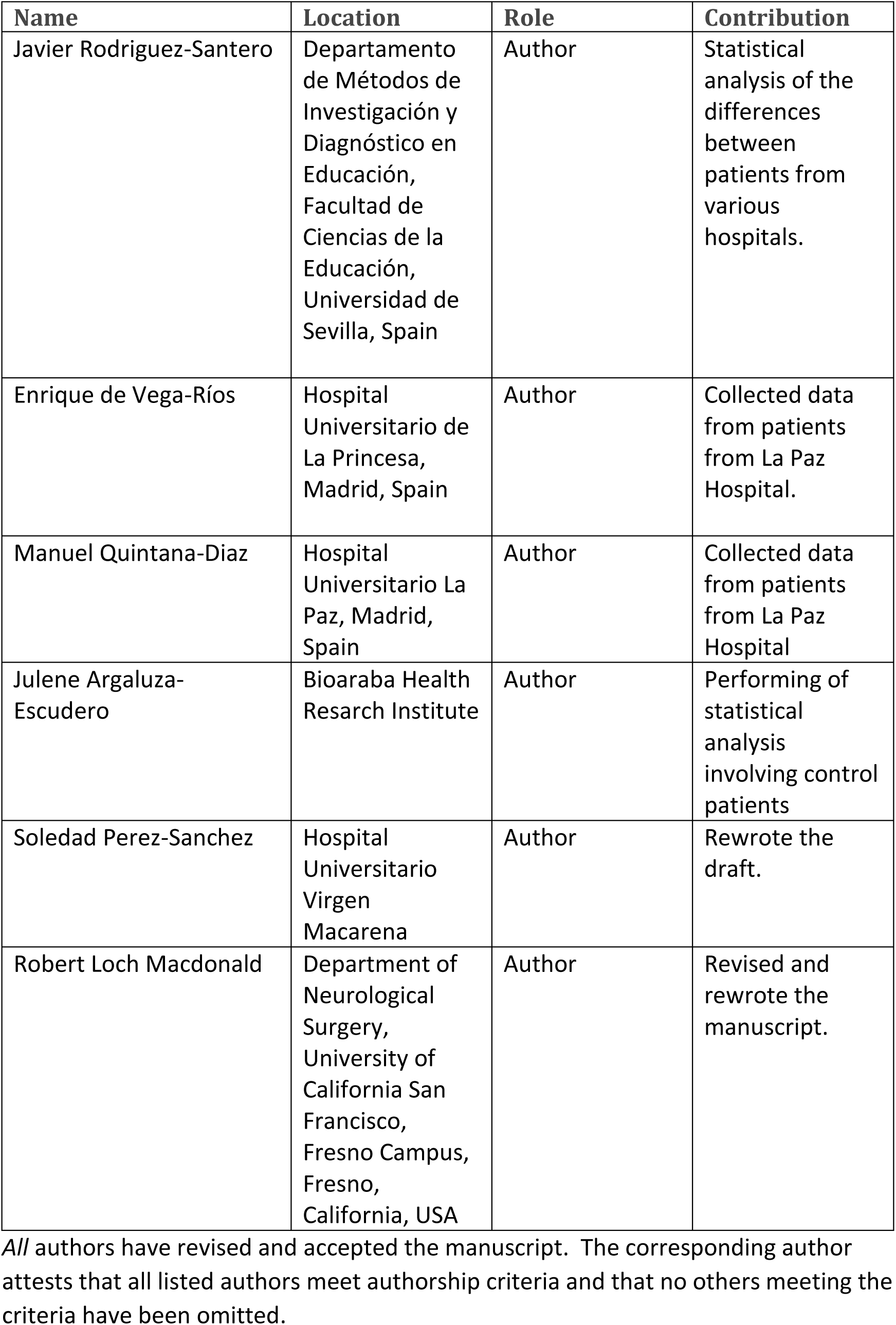

### Tripod checklist

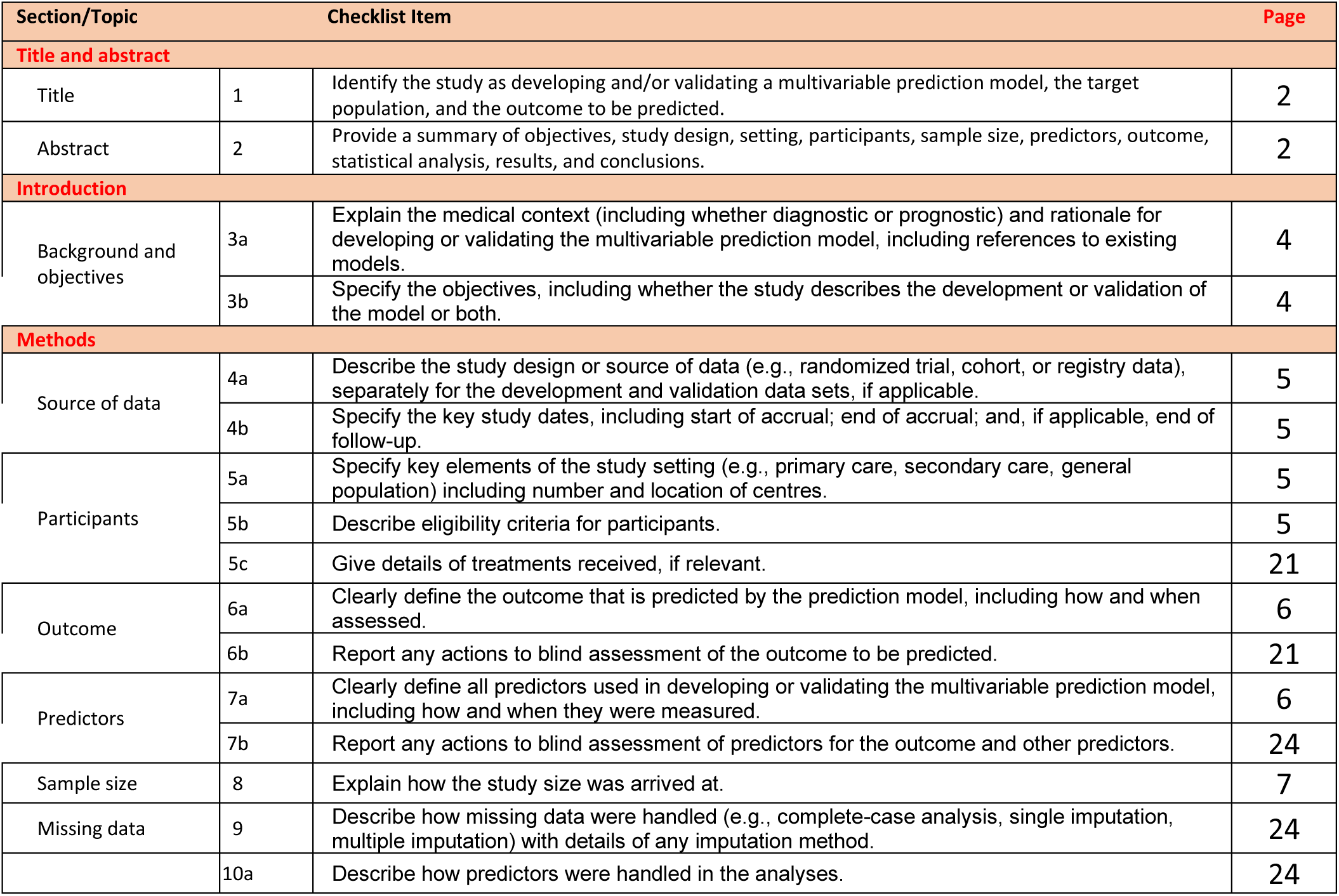
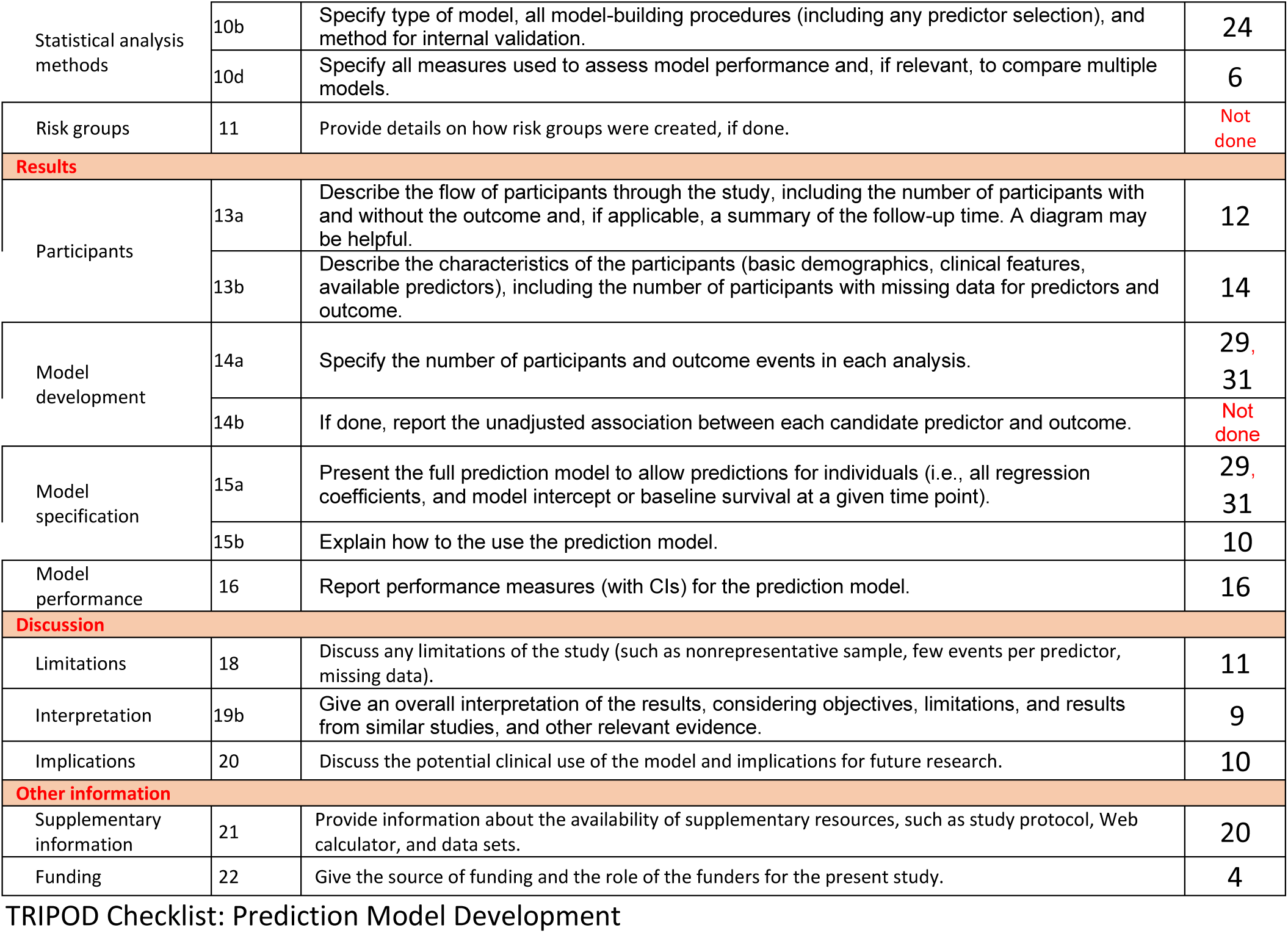

| Section/Topic |  | Checklist Item | Page |
| --- | --- | --- | --- |
| **Title and abstract** |  |  |  |
| Title | 1 | Identify the study as developing and/or validating a multivariable prediction model, the target population, and the outcome to be predicted. | 2 |
| Abstract | 2 | Provide a summary of objectives, study design, setting, participants, sample size, predictors, outcome, statistical analysis, results, and conclusions. | 2 |
| **Introduction** |  |  |  |
| Background and objectives | 3a | Explain the medical context (including whether diagnostic or prognostic) and rationale for developing or validating the multivariable prediction model, including references to existing models. | 4 |
|  | 3b | Specify the objectives, including whether the study describes the development or validation of the model or both. | 4 |
| **Methods** |  |  |  |
| Source of data | 4a | Describe the study design or source of data (e.g., randomized trial, cohort, or registry data), separately for the development and validation data sets, if applicable. | 5 |
|  | 4b | Specify the key study dates, including start of accrual; end of accrual; and, if applicable, end of follow-up. | 5 |
| Participants | 5a | Specify key elements of the study setting (e.g., primary care, secondary care, general population) including number and location of centres. | 5 |
|  | 5b | Describe eligibility criteria for participants. | 5 |
|  | 5c | Give details of treatments received, if relevant. | 21 |
| Outcome | 6a | Clearly define the outcome that is predicted by the prediction model, including how and when assessed. | 6 |
|  | 6b | Report any actions to blind assessment of the outcome to be predicted. | 21 |
| Predictors | 7a | Clearly define all predictors used in developing or validating the multivariable prediction model, including how and when they were measured. | 6 |
|  | 7b | Report any actions to blind assessment of predictors for the outcome and other predictors. | 24 |
| Sample size | 8 | Explain how the study size was arrived at. | 7 |
| Missing data | 9 | Describe how missing data were handled (e.g., complete-case analysis, single imputation, multiple imputation) with details of any imputation method. | 24 |
|  | 10a | Describe how predictors were handled in the analyses. | 24 |
| Statistical analysis methods | 10b | Specify type of model, all model-building procedures (including any predictor selection), and method for internal validation. | 24 |
|  | 10d | Specify all measures used to assess model performance and, if relevant, to compare multiple models. | 6 |
| Risk groups | 11 | Provide details on how risk groups were created, if done. | Not done |
| **Results** |  |  |  |
| Participants | 13a | Describe the flow of participants through the study, including the number of participants with and without the outcome and, if applicable, a summary of the follow-up time. A diagram may be helpful. | 12 |
|  | 13b | Describe the characteristics of the participants (basic demographics, clinical features, available predictors), including the number of participants with missing data for predictors and outcome. | 14 |
| Model development | 14a | Specify the number of participants and outcome events in each analysis. | 29, 31 |
|  | 14b | If done, report the unadjusted association between each candidate predictor and outcome. | Not done |
| Model specification | 15a | Present the full prediction model to allow predictions for individuals (i.e., all regression coefficients, and model intercept or baseline survival at a given time point). | 29, 31 |
|  | 15b | Explain how to use the prediction model. | 10 |
| Model performance | 16 | Report performance measures (with CIs) for the prediction model. | 16 |
| **Discussion** |  |  |  |
| Limitations | 18 | Discuss any limitations of the study (such as nonrepresentative sample, few events per predictor, missing data). | 11 |
| Interpretation | 19b | Give an overall interpretation of the results, considering objectives, limitations, and results from similar studies, and other relevant evidence. | 9 |
| Implications | 20 | Discuss the potential clinical use of the model and implications for future research. | 10 |
| **Other information** |  |  |  |
| Supplementary information | 21 | Provide information about the availability of supplementary resources, such as study protocol, Web calculator, and data sets. | 20 |
| Funding | 22 | Give the source of funding and the role of the funders for the present study. | 4 |

We recommend using the TRIPOD Checklist in conjunction with the TRIPOD Explanation and Elaboration document.

### STROBE Statement

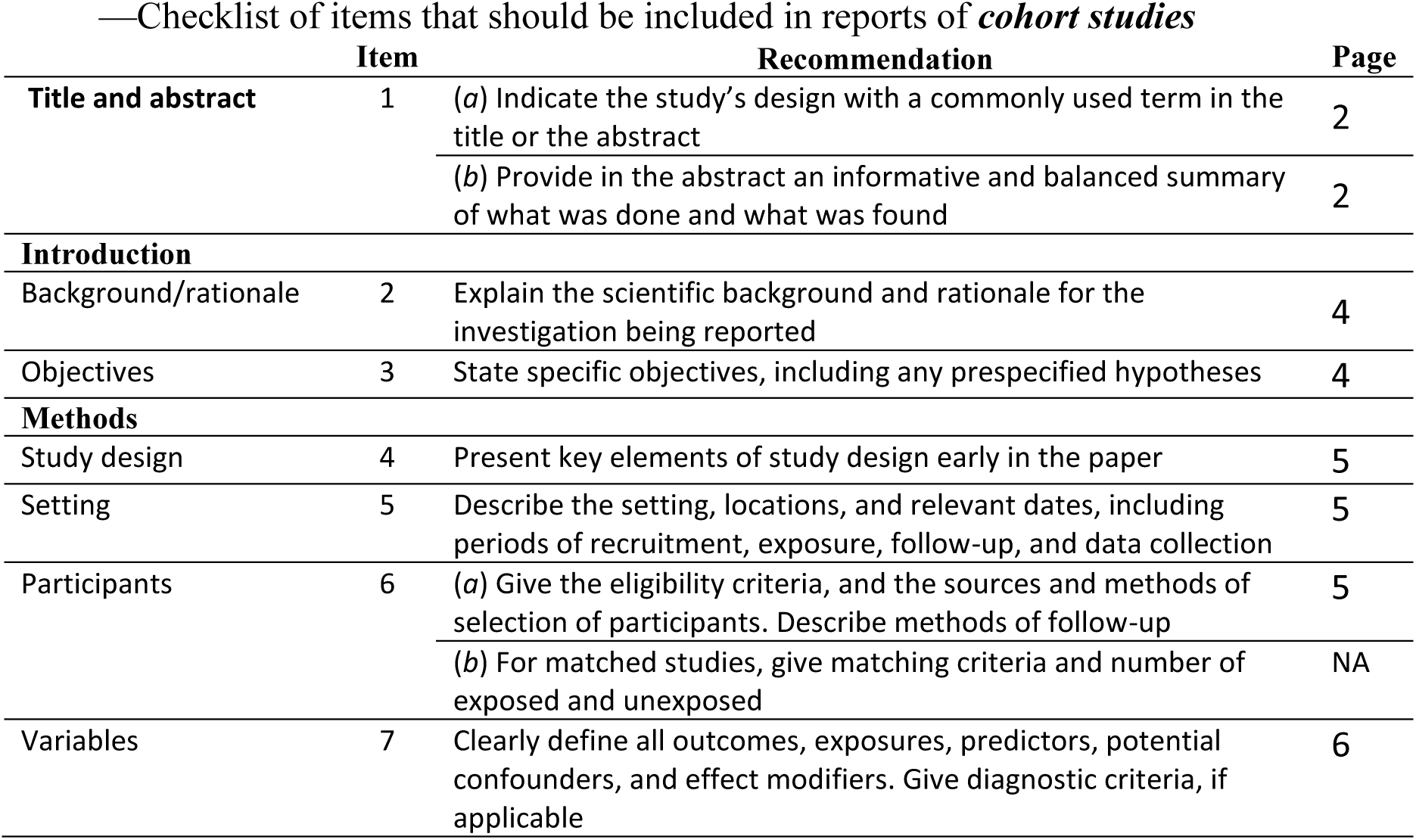
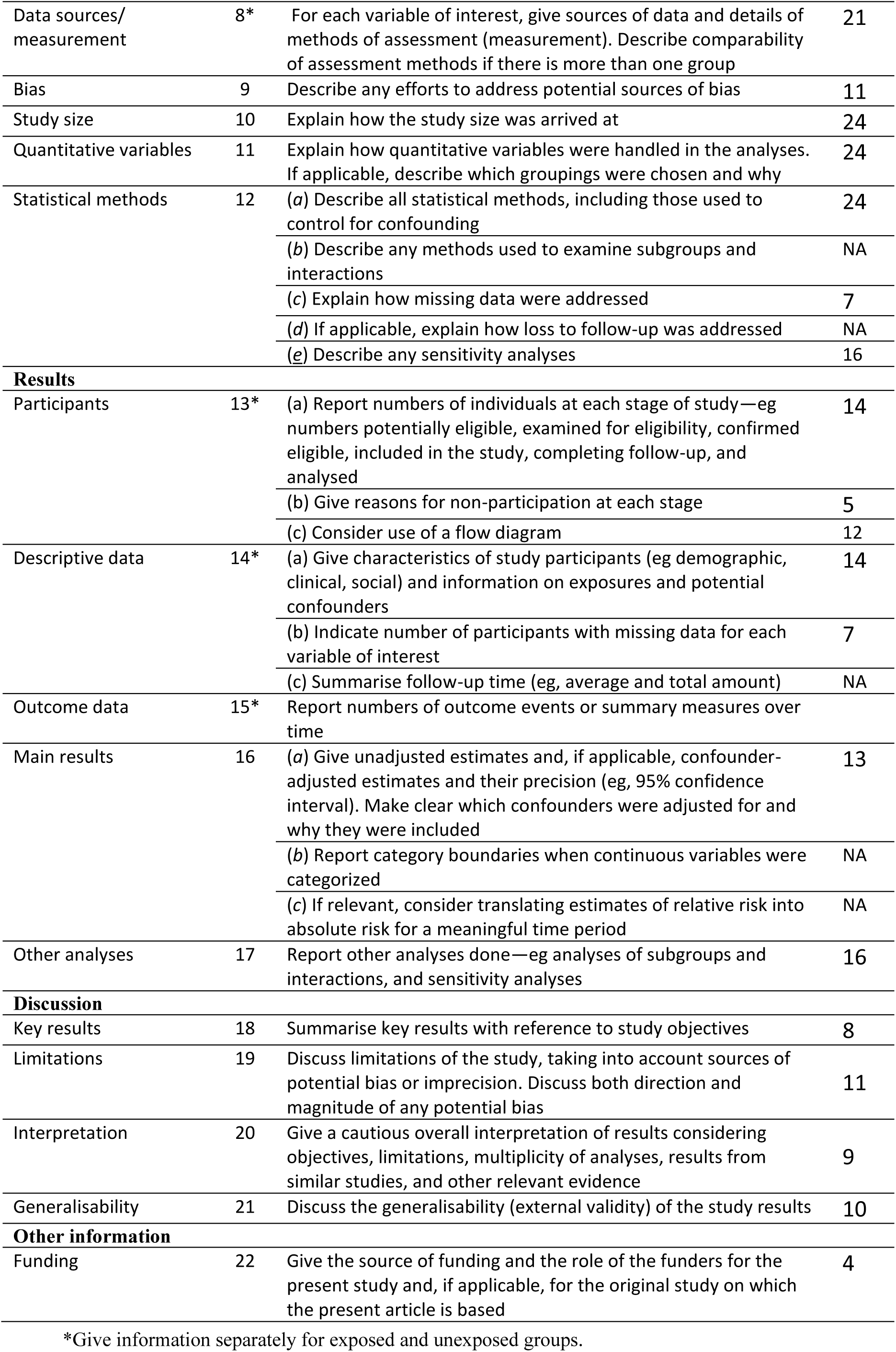
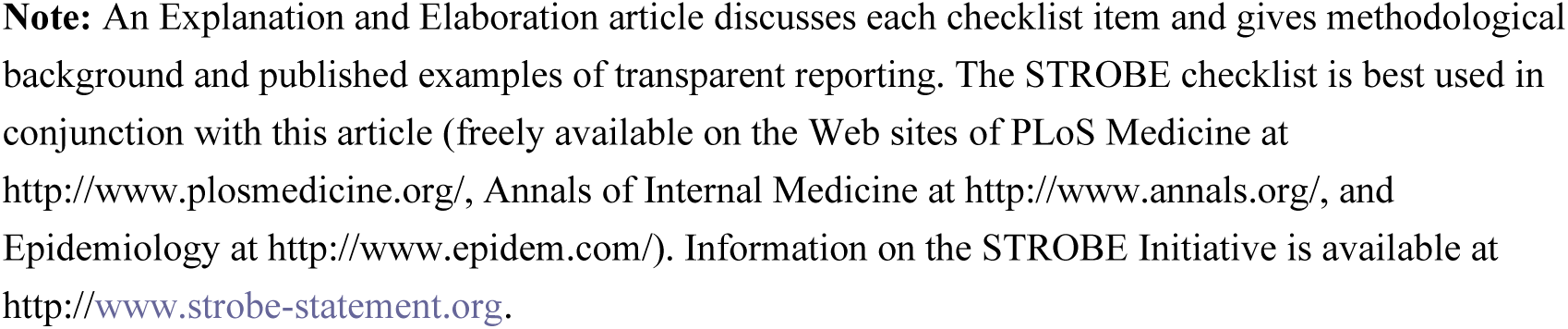

|  | Item | Recommendation | Page |
| --- | --- | --- | --- |
| **Title and abstract** | 1 | (a) Indicate the study's design with a commonly used term in the title or the abstract | 2 |
|  |  | (b) Provide in the abstract an informative and balanced summary of what was done and what was found | 2 |
| **Introduction** |  |  |  |
| Background/rationale | 2 | Explain the scientific background and rationale for the investigation being reported | 4 |
| Objectives | 3 | State specific objectives, including any prespecified hypotheses | 4 |
| **Methods** |  |  |  |
| Study design | 4 | Present key elements of study design early in the paper | 5 |
| Setting | 5 | Describe the setting, locations, and relevant dates, including periods of recruitment, exposure, follow-up, and data collection | 5 |
| Participants | 6 | (a) Give the eligibility criteria, and the sources and methods of selection of participants. Describe methods of follow-up | 5 |
|  |  | (b) For matched studies, give matching criteria and number of exposed and unexposed | NA |
| Variables | 7 | Clearly define all outcomes, exposures, predictors, potential confounders, and effect modifiers. Give diagnostic criteria, if applicable | 6 |
| Data sources/<br>measurement | 8* | For each variable of interest, give sources of data and details of methods of assessment (measurement). Describe comparability of assessment methods if there is more than one group | 21 |
| Bias | 9 | Describe any efforts to address potential sources of bias | 11 |
| Study size | 10 | Explain how the study size was arrived at | 24 |
| Quantitative variables | 11 | Explain how quantitative variables were handled in the analyses. If applicable, describe which groupings were chosen and why | 24 |
| Statistical methods | 12 | (a) Describe all statistical methods, including those used to control for confounding | 24 |
|  |  | (b) Describe any methods used to examine subgroups and interactions | NA |
|  |  | (c) Explain how missing data were addressed | 7 |
|  |  | (d) If applicable, explain how loss to follow-up was addressed | NA |
|  |  | (e) Describe any sensitivity analyses | 16 |
| **Results** |  |  |  |
| Participants | 13* | (a) Report numbers of individuals at each stage of study—eg numbers potentially eligible, examined for eligibility, confirmed eligible, included in the study, completing follow-up, and analysed | 14 |
|  |  | (b) Give reasons for non-participation at each stage | 5 |
|  |  | (c) Consider use of a flow diagram | 12 |
| Descriptive data | 14* | (a) Give characteristics of study participants (eg demographic, clinical, social) and information on exposures and potential confounders | 14 |
|  |  | (b) Indicate number of participants with missing data for each variable of interest | 7 |
|  |  | (c) Summarise follow-up time (eg, average and total amount) | NA |
| Outcome data | 15* | Report numbers of outcome events or summary measures over time |  |
| Main results | 16 | (a) Give unadjusted estimates and, if applicable, confounder-adjusted estimates and their precision (eg, 95% confidence interval). Make clear which confounders were adjusted for and why they were included | 13 |
|  |  | (b) Report category boundaries when continuous variables were categorized | NA |
|  |  | (c) If relevant, consider translating estimates of relative risk into absolute risk for a meaningful time period | NA |
| Other analyses | 17 | Report other analyses done—eg analyses of subgroups and interactions, and sensitivity analyses | 16 |
| **Discussion** |  |  |  |
| Key results | 18 | Summarise key results with reference to study objectives | 8 |
| Limitations | 19 | Discuss limitations of the study, taking into account sources of potential bias or imprecision. Discuss both direction and magnitude of any potential bias | 11 |
| Interpretation | 20 | Give a cautious overall interpretation of results considering objectives, limitations, multiplicity of analyses, results from similar studies, and other relevant evidence | 9 |
| Generalisability | 21 | Discuss the generalisability (external validity) of the study results | 10 |
| **Other information** |  |  |  |
| Funding | 22 | Give the source of funding and the role of the funders for the present study and, if applicable, for the original study on which the present article is based | 4 |

\*Give information separately for exposed and unexposed groups.
**Note:** An Explanation and Elaboration article discusses each checklist item and gives methodological background and published examples of transparent reporting. The STROBE checklist is best used in conjunction with this article (freely available on the Web sites of PLoS Medicine at http://www.plosmedicine.org/, Annals of Internal Medicine at http://www.annals.org/, and Epidemiology at http://www.epidem.com/). Information on the STROBE Initiative is available at http://www.strobe-statement.org.

## Notes

### Competing Interest Statement

Juan Jose Egea-Guerrero and Marcin Balcerzyk are inventors of the patent EPO EP3664027A1 and USPTO patent US11344260B2 to the described biomarker panel method. Marcin Balcerzyk is an owner of a company CT Provision SLU located in Spain. R. Loch Macdonald discloses receiving consulting fees from CSL Behring, LLC, Acasti Pharma and Idorsia Pharmaceuticals. Gabriel Cepeda-Carrion, Angel Parrado-Gallego, Zaida Ruiz de Azua Lopez, Angel Vilches-Arenas, Javier Rodriguez-Santero, Enrique de Vega-Rios, Manuel Quintana-Diaz, Julene Argaluza-Escudero, Soledad Perez-Sanchez report no disclosures. The study is not industry sponsored. The patients for this study were recruited thanks to a grant from Consejeria de Igualdad, Salud y Politicas Sociales de Andalucia, Spain (PI-0136-2012).

### Funding Statement

1. The study is not industry sponsored. The patients for this study were recruited thanks to a grant from Consejeria de Igualdad, Salud y Politicas Sociales de Andalucia, Spain (PI-0136-2012). 2. The study was partially funded by the project CI19-00068 from CaixaImpulse program of La Caixa Foundation (Barcelona, Spain).

### Author Declarations

This research project was overseen and approved by Hospital Universitario Virgen del Rocio in Seville, Spain hospital ethics committee (Cod. CEI2012PI/228). Informed consent was obtained from all patients participating in the study or from their close relatives.

### Summary of Updates

The affiliations of Marcin Balcerzyk were corrected.

## References

1. Rorden C, Bonilha L, Fridriksson J, et al. Age-specific CT and MRI templates for spatial normalization. NeuroImage 2012;61(4):957–65. doi: 10.1016/j.neuroimage.2012.03.020 [published Online First: 2012/03/24]

2. Etminan N, Chang H-S, Hackenberg K, et al. Worldwide Incidence of Aneurysmal Subarachnoid Hemorrhage According to Region, Time Period, Blood Pressure, and Smoking Prevalence in the Population A Systematic Review and Meta-analysis. Jama Neurology 2019;76(5):588–97. doi: 10.1001/jamaneurol.2019.0006

3. Fisher CM, Kistler JP, Davis JM. RELATION OF CEREBRAL VASOSPASM TO SUBARACHNOID HEMORRHAGE VISUALIZED BY COMPUTERIZED TOMOGRAPHIC SCANNING. Neurosurgery 1980;6(1):1–9. doi: 10.1227/00006123-198001000-00001

4. Claassen J, Bernardini GL, Kreiter K, et al. Effect of cisternal and ventricular blood on risk of delayed cerebral ischemia after subarachnoid hemorrhage - The Fisher scale revisited. Stroke 2001;32(9):2012–20. doi: 10.1161/hs0901.095677

5. Hijdra A, van Gijn J, Nagelkerke NJ, et al. Prediction of delayed cerebral ischemia, rebleeding, and outcome after aneurysmal subarachnoid hemorrhage. Stroke 1988;19(10):1250–6.

6. Vergouwen MD, Vermeulen M, van Gijn J, et al. Definition of delayed cerebral ischemia after aneurysmal subarachnoid hemorrhage as an outcome event in clinical trials and observational studies: proposal of a multidisciplinary research group. Stroke; a journal of cerebral circulation 2010;41(10):2391–95. doi: 10.1161/STROKEAHA.110.589275

7. Edlow JA, Caplan LR. Avoiding Pitfalls in the Diagnosis of Subarachnoid Hemorrhage. New England Journal of Medicine 2000;342(1):29–36. doi: doi:10.1056/NEJM200001063420106

8. Zhang X, Hong H, Wang X, et al. Serum Gas6 contributes to clinical outcome after aneurysmal subarachnoid hemorrhage: A prospective cohort study. Clinica Chimica Acta 2022;533:96–103. doi: 10.1016/j.cca.2022.06.016

9. Tatli O, Yadigaroglu M, Demir S, et al. The value of glial fibrillary acidic protein levels in the diagnosis and prognosis of subarachnoid hemorrhage. Hong Kong J Emerg Med 2022;29(3):151–60. doi: 10.1177/1024907920915054

10. Hathidara MY, Campos Y, Chandrashekhar S, et al. Scoring system to predict hospital outcome after subarachnoid hemorrhage–incorporating systemic response: The CRIG score. Journal of Stroke and Cerebrovascular Diseases 2022;31(8) doi: 10.1016/j.jstrokecerebrovasdis.2022.106577

11. Xie B, Lin Y, Wu X, et al. Reduced Admission Serum Fibrinogen Levels Predict 6-Month Mortality of Poor-Grade Aneurysmal Subarachnoid Hemorrhage. World Neurosurgery 2020;136:e24–e32. doi: 10.1016/j.wneu.2019.08.155

12. Ding CY, Cai HP, Ge HL, et al. Is Admission Lipoprotein-Associated Phospholipase A2 a Novel Predictor of Vasospasm and Outcome in Patients with Aneurysmal Subarachnoid Hemorrhage? Neurosurgery 2020;86(1):122–30. doi: 10.1093/neuros/nyz041

13. Bacigaluppi S, Ivaldi F, Bragazzi NL, et al. An Early Increase of Blood Leukocyte Subsets in Aneurysmal Subarachnoid Hemorrhage Is Predictive of Vasospasm. Frontiers in Neurology 2020;11 doi: 10.3389/fneur.2020.587039

14. Zhang D, Zhuang Z, Wei Y, et al. Association of Admission Serum Glucose– Phosphate Ratio with Severity and Prognosis of Aneurysmal Subarachnoid Hemorrhage. World Neurosurgery 2019;127:e1145–e51. doi: 10.1016/j.wneu.2019.04.071

15. Zhang D, Yan H, Wei Y, et al. C-Reactive Protein/Albumin Ratio Correlates With Disease Severity and Predicts Outcome in Patients With Aneurysmal Subarachnoid Hemorrhage. Frontiers in Neurology 2019;10 doi: 10.3389/fneur.2019.01186

16. Cai H, Zheng S, Cai B, et al. Neuroglobin as a Novel Biomarker for Predicting Poor Outcomes in Aneurysmal Subarachnoid Hemorrhage. World Neurosurgery 2018;116:e258–e65. doi: 10.1016/j.wneu.2018.04.184

17. Liu H, Liu Y, Zhao J, et al. Prognostic value of plasma galectin-3 levels after aneurysmal subarachnoid hemorrhage. Brain Behav 2016;6(10) doi: 10.1002/brb3.543

18. De Oliveira Manoel AL, Jaja BN, Germans MR, et al. The VASOGRADE: A Simple Grading Scale for Prediction of Delayed Cerebral Ischemia after Subarachnoid Hemorrhage. Stroke 2015;46(7):1826–31. doi: 10.1161/STROKEAHA.115.008728

19. Lagares A, Jiménez-Roldán L, Gomez PA, et al. Prognostic Value of the Amount of Bleeding After Aneurysmal Subarachnoid Hemorrhage: A Quantitative Volumetric Study. Neurosurgery 2015;77(6):898–906. doi: 10.1227/NEU.0000000000000927

20. Lee H, Perry JJ, English SW, et al. Clinical prediction of delayed cerebral ischemia in aneurysmal subarachnoid hemorrhage. J Neurosurg 2019;130(6):1914–21. doi: 10.3171/2018.1.jns172715

21. Hickmann AK, Langner S, Kirsch M, et al. The value of perfusion computed tomography in predicting clinically relevant vasospasm in patients with aneurysmal subarachnoid hemorrhage. Neurosurgical Review 2013;36(2):267–78. doi: 10.1007/s10143-012-0430-1

22. Turck N, Vutskits L, Sanchez-Pena P, et al. A multiparameter panel method for outcome prediction following aneurysmal subarachnoid hemorrhage. Intensive Care Medicine 2010;36(1):107–15. doi: 10.1007/s00134-009-1641-y

23. Rodríguez-Rodríguez A, Egea-Guerrero JJ, Ruiz De Azúa-López Z, et al. Biomarkers of vasospasm development and outcome in aneurysmal subarachnoid hemorrhage. Journal of the Neurological Sciences 2014;341(1-2):119–27. doi: 10.1016/j.jns.2014.04.020

24. Jimenez-Roldan L, Alen JF, Gomez PA, et al. Volumetric analysis of subarachnoid hemorrhage: assessment of the reliability of two computerized methods and their comparison with other radiographic scales. J Neurosurg 2013;118(1):84–93. doi: 10.3171/2012.8.JNS12100 [published Online First: 2012/09/25]

25. Prakash B, Hu J, Morgan TC, et al. Comparison of 3-Segmentation Techniques for Intraventricular and Intracerebral Hemorrhages in Unenhanced Computed Tomography Scans. Journal of Computer Assisted Tomography 2012;36(1):109–20. doi: 10.1097/RCT.0b013e318245c1fa

26. Prakash KNB, Zhou S, Morgan TC, et al. Segmentation and quantification of intra-ventricular/cerebral hemorrhage in CT scans by modified distance regularized level set evolution technique. International Journal of Computer Assisted Radiology and Surgery 2012;7(5):785–98. doi: 10.1007/s11548-012-0670-0

27. Ko SB, Choi HA, Carpenter AM, et al. Quantitative analysis of hemorrhage volume for predicting delayed cerebral ischemia after subarachnoid hemorrhage. Stroke 2011;42(3):669–74. doi: 10.1161/STROKEAHA.110.600775

28. Kothari RU, Brott T, Broderick JP, et al. The ABCs of measuring intracerebral hemorrhage volumes. Stroke 1996;27(8):1304–05. doi: 10.1161/01.STR.27.8.1304

29. Kosior JC, Idris S, Dowlatshahi D, et al. Quantomo: validation of a computer-assisted methodology for the volumetric analysis of intracerebral haemorrhage. International Journal of Stroke 2011;6(4):302–05. doi: 10.1111/j.1747-4949.2010.00579.x

30. von Elm E, Altman DG, Egger M, et al. The Strengthening the Reporting of Observational Studies in Epidemiology (STROBE) statement: guidelines for reporting observational studies. The Lancet 2007;370(9596):1453–57. doi: 10.1016/S0140-6736(07)61602-X

31. Burke HB. TRansparent reporting of a multivariable prediction model for individual prognosis or diagnosis (tripod). Annals of Internal Medicine 2015;162(10):735. doi: 10.7326/L15-5093

32. William E. Hunt, Robert M. Hess. Surgical Risk as Related to Time of Intervention in the Repair of Intracranial Aneurysms. J Neurosurg 1968;28(1):14–20. doi: 10.3171/jns.1968.28.1.0014

33. Teasdale GM, Drake CG, Hunt W, et al. A universal subarachnoid hemorrhage scale: report of a committee of the World Federation of Neurosurgical Societies. J Neurol Neurosurg Psychiatry 1988;51(11):1457. doi: 10.1136/jnnp.51.11.1457

34. PMOD Biomedical Image Quantification [program]. 4.3 version. Zurich, Switzerland: PMOD Technologies Ltd, 2018.

35. Robin X, Turck N, Hainard A, et al. pROC: an open-source package for R and S+ to analyze and compare ROC curves. BMC Bioinformatics 2011;12(1):1–8. doi: 10.1186/1471-2105-12-77

36. Ramos LA, van der Steen WE, Sales Barros R, et al. Machine learning improves prediction of delayed cerebral ischemia in patients with subarachnoid hemorrhage. J Neurointerv Surg 2019;11(5):497–502. doi: 10.1136/neurintsurg-2018-014258 [published Online First: 20181110]

37. Jaja BNR, Cusimano MD, Etminan N, et al. Clinical prediction models for aneurysmal subarachnoid hemorrhage: A systematic review. Neurocritical Care 2013;18(1):143–53. doi: 10.1007/s12028-012-9792-z

38. Chaudhry SR, Guresir A, Stoffel-Wagner B, et al. Systemic High-Mobility Group Box-1: A Novel Predictive Biomarker for Cerebral Vasospasm in Aneurysmal Subarachnoid Hemorrhage. Crit Care Med 2018 doi: 10.1097/CCM.0000000000003319 [published Online First: 2018/07/22]

39. Oertel M, Schumacher U, McArthur DL, et al. S-100B and NSE: markers of initial impact of subarachnoid haemorrhage and their relation to vasospasm and outcome. Journal of Clinical Neuroscience 2006;13(8):834–40. doi: 10.1016/j.jocn.2005.11.030

40. Turck N, Vutskits L, Sanchez-Pena P, et al. A multiparameter panel method for outcome prediction following aneurysmal subarachnoid hemorrhage. Intensive Care Medicine 2009;36(1):107–15. doi: 10.1007/s00134-009-1641-y

41. Lindegaard KF, Nornes H, Bakke SJ, et al. Cerebral vasospasm diagnosis by means of angiography and blood velocity measurements. Acta Neurochir 1989;100(1-2):12–24. doi: 10.1007/bf01405268

42. Snider SB, Migdady I, Larose SL, et al. Transcranial-Doppler-Measured Vasospasm Severity is Associated with Delayed Cerebral Infarction After Subarachnoid Hemorrhage. Neurocritical Care 2022;36(3):815–21. doi: 10.1007/s12028-021-01382-2

43. Narayanan R, El-Sayed MA. Effect of Catalysis on the Stability of Metallic Nanoparticles: Suzuki Reaction Catalyzed by PVP-Palladium Nanoparticles. J Am Chem Soc 2003;125(27):8340–47. doi: 10.1021/ja035044x

